# Machine Learning Models for the Prediction of Early-Onset Bipolar Using Electronic Health Records

**DOI:** 10.1101/2024.02.19.24302919

**Authors:** Bo Wang, Yi-Han Sheu, Hyunjoon Lee, Robert G. Mealer, Victor M. Castro, Jordan W. Smoller

## Abstract

**Objective:** Early identification of bipolar disorder (BD) provides an important opportunity for timely intervention. In this study, we aimed to develop machine learning models using large-scale electronic health record (EHR) data including clinical notes for predicting early-onset BD.

**Method:** Structured and unstructured data were extracted from the longitudinal EHR of the Mass General Brigham health system. We defined three cohorts aged 10 – 25 years: (1) the full youth cohort (N=300,398); (2) a sub-cohort defined by having a mental health visit (N=105,461); (3) a sub-cohort defined by having a diagnosis of mood disorder or ADHD (N=35,213). By adopting a prospective landmark modeling approach that aligns with clinical practice, we developed and validated a range of machine learning models including neural network-based models, across different cohorts and prediction windows.

**Results:** We found the two tree-based models, Random forests (RF) and light gradient-boosting machine (LGBM), achieving good discriminative performance across different clinical settings (area under the receiver operating characteristic curve 0.76-0.88 for RF and 0.74-0.89 for LGBM). In addition, we showed comparable performance can be achieved with a greatly reduced set of features, demonstrating computational efficiency can be attained without significant compromise of model accuracy.

**Conclusion:** Good discriminative performance for early-onset BD is achieved utilizing large-scale EHR data. Our study offers a scalable and accurate method for identifying youth at risk for BD that could help inform clinical decision making and facilitate early intervention. Future work includes evaluating the portability of our approach to other healthcare systems and exploring considerations regarding possible implementation.

## 1. Introduction

Bipolar disorder (BD) is a serious mental illness typically characterized by recurrent episodes of mania (or hypomania) and depression^1^. A recent analysis of data from 29 countries revealed a lifetime prevalence of 2.3% (among females) - 2.5% (among males) and a median age of onset of approximately 20 years^2^. Earlier-onset BD has been associated with a more severe course including higher rates of recurrent mood episodes, comorbid psychopathology, and suicide attempts^3–6^. Unfortunately, the interval between onset of the disorder and accurate diagnosis or treatment averages 6-10 years, and longer duration of untreated BD is associated with poorer outcomes^7–11^. Given this, early identification of BD provides an important opportunity for intervention and prevention of adverse sequelae.

A number of clinical indicators have been reported to be associated with risk for BD, including positive family history, early-onset depressive episodes, and history of attention deficit/hyperactivity disorder (ADHD)^12^. However, the utility of these indicators for individualized risk stratification has not been demonstrated. Several studies have used longitudinal assessments of children at risk for BD to predict incident disorder. In a community-based longitudinal cohort of offspring (N = 412 under age 18) of parents with BD, Hafeman and colleagues^13^ developed a Cox proportional hazards model to predict conversion to a bipolar spectrum disorder and achieved good discrimination at 5 years (AUC = 0.76). Uchida and colleagues^14^ applied machine learning to longitudinal pediatric cohorts (N = 492) ascertained for studies of ADHD. In that study, a random forest model incorporating a battery of survey instruments achieved an AUC of 0.75 for predicting conversion to bipolar I disorder over 10 years of follow-up. However, both studies comprised relatively small samples (with fewer than 55 bipolar outcomes in each) and relied on diagnostic interviews and survey data that may not be readily available outside research protocols.

In recent years, the widespread availability of large-scale electronic health record (EHR) data has provided an important resource for prediction or detection of medical and psychiatric outcomes. Machine learning methods are well-suited to leverage these high dimensional data and have been used successfully in the prediction of a range of mental health outcomes including suicide-related behavior, psychosis, and treatment response^15–20^. A recent study by the PsycheMERGE consortium validated EHR-based machine learning models to predict BD across three healthcare systems. The best-performing models achieved high discrimination (AUCs 0.82 – 0.87) and patients in the top 1% of predicted risk had up to a 19-fold increased rate of BD compared to base rates. Importantly, however, this study was restricted to adult patients. Here, we develop and validate machine learning models with the aim of predicting early-onset BD among youth aged 10 – 25 years in a large healthcare system. This work represents to our knowledge the first such effort and extends prior work by 1) incorporating natural language processing (NLP) of narrative notes; 2) including deep learning (neural network) models; 3) using a landmark model framework that avoids temporal bias^21^; and 4) developing models for the full cohort as well as two sub-cohorts respectively defined by having a mental health visit or a diagnosis of mood disorder or ADHD prior to the onset of BD.

## 2. Methods

An overview of our data processing and modelling pipeline is illustrated in Figure 1. We show the comprehensive version of the data filtering and cohort building flow chart in Supplementary Figure 1, and the detailed description of feature extraction and model choices in <u>Feature</u> <u>extraction</u>and <u>Model building</u>.

**Figure 1:**
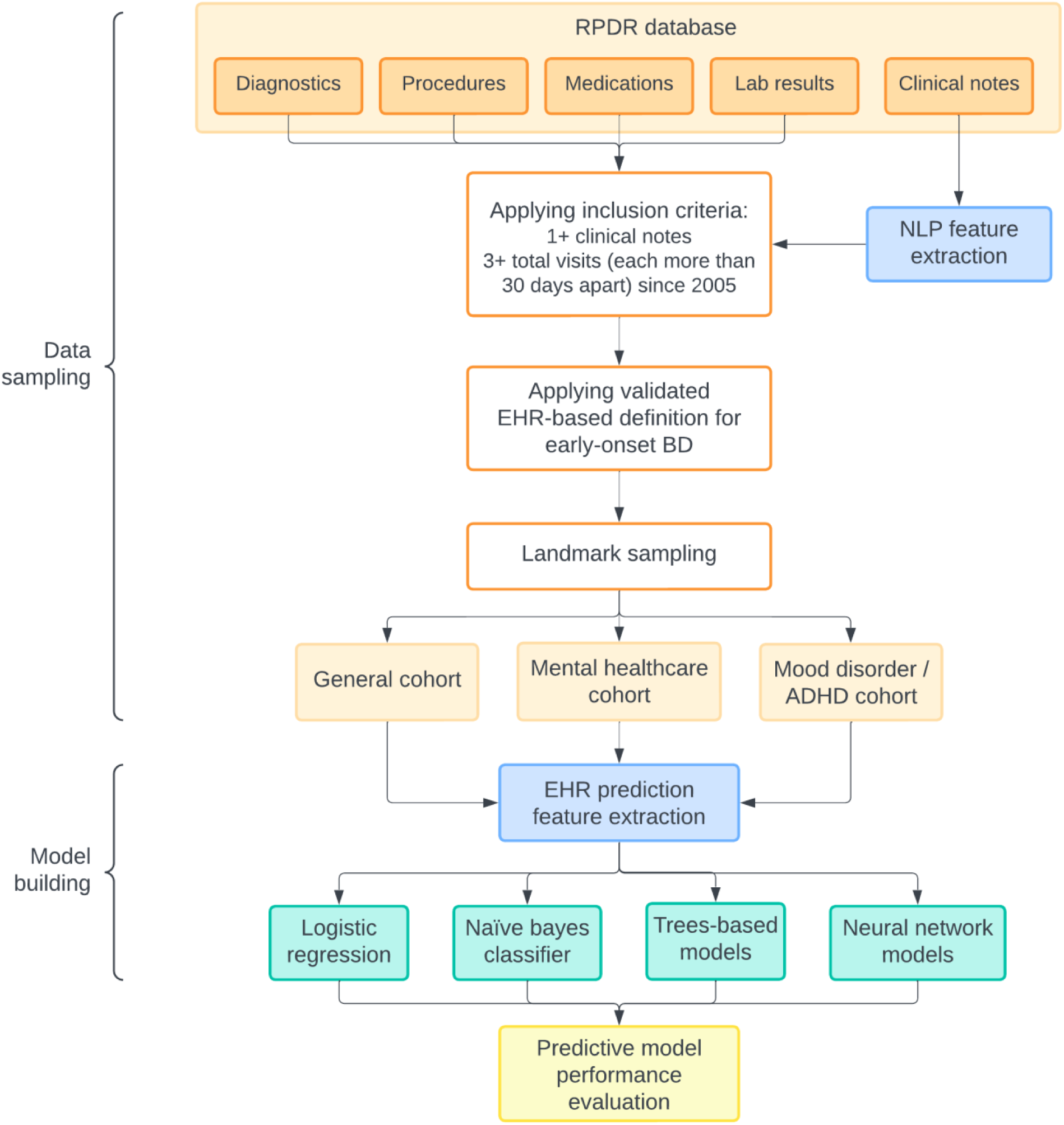
Data processing and model building flowchart, describing the major steps taken to arrive at the final datasets used for training and evaluating different models for predicting early-onset BD.

### 2.1 Data source and study population

The data for this study were obtained from the Mass General Brigham (MGB) Research Patient Data Registry (RPDR)^22^, a centralized data registry of clinical information from the EHRs across the MGB health system. The RPDR database includes approximately 7 million patients from 8 hospitals, including two major teaching hospitals, Massachusetts General Hospital and Brigham and Women’s Hospital, encompassing a wide range of patient characteristics including structured data (demographics, diagnoses, medications, procedures, and lab tests) and unstructured narrative notes. We queried the RPDR for all visits (i.e., inpatient, outpatient and emergency department visits) occurring between Dec 1997 and June 2019 who met a data floor of having at least 1 clinical note, and at least 3 visits (each more than 30 days apart) since 2005. This queried dataset was deidentified for all modelling and analyses.

### 2.2 Definition of Early Onset Bipolar Disorder

Bipolar disorder (BD) was defined using an EHR-based phenotyping algorithm referred to as “ICCBD-coded-broad” developed by International Cohort Collection for Bipolar Disorder (ICCBD)^23^. In prior work, this algorithm has been validated against in-person semi-structured diagnostic interviews (SCID-IV) by doctoral level clinicians and achieved high positive predictive value for diagnosing BD (PPV = 0.80)^23^. Cases are defined by having 1) at least two International Classification of Disease (ICD) 9/10 codes for BD AND 2) a predominance of BD diagnoses AND 3) treatment with at least two medications (mood stabilizers or antipsychotics) commonly used for BD. We defined early-onset BD as having the first BD diagnosis code onset of BD between age 10 (inclusive) and 25 (non-inclusive). They also were required to meet the full ICCBD coded-broad criteria (details in Appendix A.1). To validate this definition of early-onset BD, an experienced, board-certified psychiatrist performed chart reviews for a random sample of 99 patients determined to have early-onset BD by our definition. We adopted the ICCBD diagnostician review protocol^23^ for our chart review. The positive predictive value (PPV) of our ICCBD-coded-broad early-onset BD definition by chart review was high (89.9%) in this study population (i.e., patients between ages 10 and 25). We thus applied this definition of early-onset BD in our cohort building.

### 2.3 Cohort building

#### 2.3.1 Patient eligibility

We defined three patient cohorts of interest who are at risk of early-onset BD within the RDPR by clinical settings: (1) a “general cohort”, which comprised the full study population; (2) a “mental healthcare cohort”, applying the same condition as the general cohort, but requiring at least one mental health visit (defined in Appendix A.2) before age 25 and (for those who meet our definition of early-onset BD) before their first BD diagnosis; and (3) a “mood disorder/ADHD cohort”, applying the same condition as the general cohort, but requiring at least one non-bipolar mood disorder or ADHD diagnosis (defined in Appendix A.3) before age 25 and (for those who meet our definition of early-onset BD) before their first BD diagnosis. The mental healthcare and mood disorder/ADHD cohorts were designed to address common clinical scenarios in which a prediction algorithm might be especially useful.

#### 2.3.2 Landmark sampling

Supervised learning prediction models are commonly trained using case-control sampling. However, this design does not mimic real-world practice, where a clinician would not have access to the (future) case vs. control status of a given individual. This mismatch has been shown to induce a “temporal bias” that can result in spurious inflation of effect sizes and render the model unsuitable for prospective prediction^21^. An alternative approach, used here, avoids this problem by designating a time point of interest (the “landmark time”) from which prospective prediction is made without pre-specifying subsequent case or control status^24–27^. Data used for prediction are then sampled prior to the landmark time and predictions of the outcome of interest are made for pre-defined prediction windows (e.g. over the next year).

For each of the three cohorts defined above, we randomly sampled one visit per patient between ages 10 and 25, which we designated the “landmark visit” (i.e., the visit from which a prediction is made). We required at least 2 visits and a minimum of 365 days of EHR data prior to the landmark visit. For the patients who later developed early-onset BD, we sampled the “landmark visit” after the first two visits and before their first BD diagnosis. To avoid potential information leakage, we also required that the landmark visit occur before any visit containing BD-related mentions in narrative notes (by NLP). Figure 2 shows a patient timeline for the prediction task using the RPDR database.

**Figure 2:**
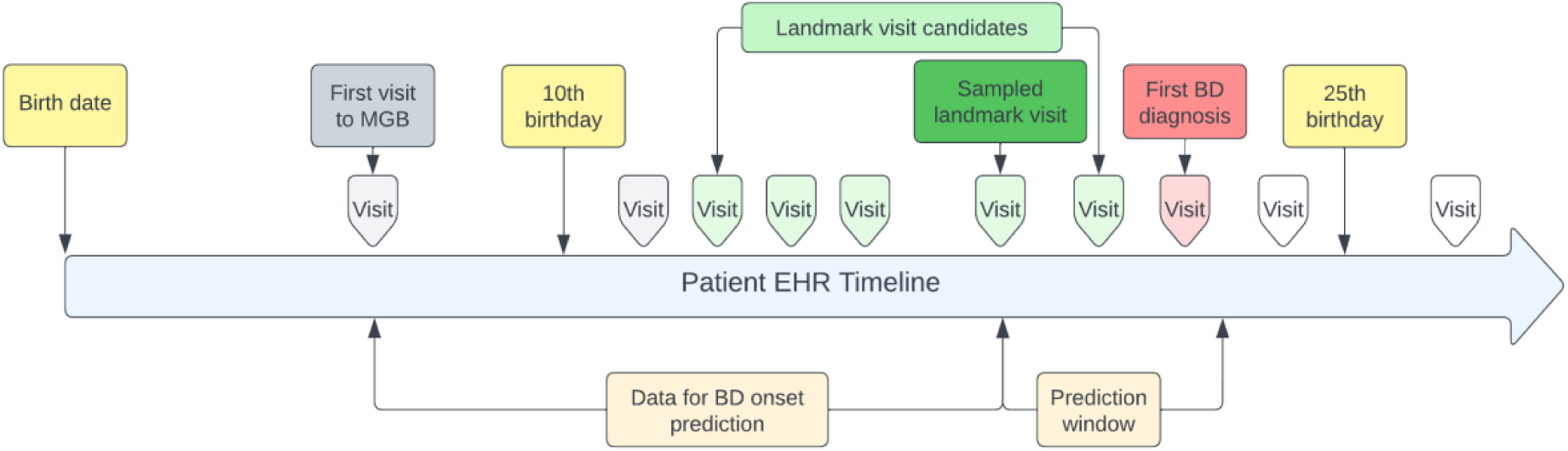
Patient EHR timeline including the sampled landmark visit, the observation period and prediction window used for the prediction tasks.

#### 2.3.3 Observation period and prediction window

After the landmark visit is sampled, each patient’s EHR timeline is divided into two segments: 1), the historical data used as predictors for BD onset prediction (i.e., the observation period), comprising all data up to and including the landmark visit for each patient; 2), the “prediction window”: the time window looking forward from the landmark visit, during which onset of BD was assessed. We examined 4 different prediction windows following the landmark visit, namely 6 months, 1 year, 2 years, and 3 years.

### 2.4 Feature extraction

The prediction features for each patient were derived using both structured EHR data and unstructured clinical notes prior to and including the landmark visit. Among the structured data, we extracted CPT-4 codes^28^ from the procedure data, RXNORM codes^29^ from the medication data, and converted ICD-9-CM and ICD-10-CM codes^30,31^ to PheWAS codes (“phecodes”) (https://phewascatalog.org/phecodes) that have been shown to capture clinically meaningful concepts^32^. We also used LOINC codes^33^ from the laboratory data and merged them with a variable representing the recorded outcome of a given lab value as a categorical variable (high, low, abnormal, normal and undetermined). In addition, using a rule-based NLP system previously developed for enhancing psychiatric research^34,35^ (described in Appendix B), we identified the presence of any mental health related concepts at each visit, and further tagged concepts as negated (NEG), family history mention (FH), or negated family history (NFH). Overall, we extracted and used 1488 mental health related concepts from all the clinical notes, which we refer to as NLP CUIs (i.e., concept unique identifiers^36^).

We removed rare codes and NLP CUIs (occurring in fewer than 10 patients in each of the study cohorts) and encoded all aforementioned features using “bag-of-codes” representation^37,38^. Lastly, we included demographic (i.e., age, gender, race, ethnicity) and health utilization (i.e. visit counts, number of ICD codes, number of procedure codes and time since the first ICD code, all computed at the landmark visit) features. Apart from the attention-based neural network models, each feature is scaled by its maximum absolute value prior to model training.

### 2.5 Model building and evaluation

We benchmarked 7 machine learning models, covering a range of architecture and complexity, on the 3 study cohorts (i.e., “general cohort”, “mental healthcare cohort”, and “mood disorder/ADHD cohort”) each with 4 risk prediction windows (i.e., 6 months, 1 year, 2 years and 3 years):

- Regularized logistic regression (LR) and Naïve Bayes classifier (NBC) are commonly used as baseline models in EHR-based prediction studies^18,39–42^.
- Random forests (RF) and LightGBM^43^ (LGBM) are two representative tree-based models that have consistently shown competitive performance across different tabular datasets^44,45^. As a result they are used for a wide range of clinical prediction tasks^14,27,41,46–48^.
- Multilayer perceptron (MLP): also known as multilayer feedforward neural network. It has been used as a baseline neural network model in various existing clinical prediction studies^38,40,49^.
- Wide & Deep Learning^50^ (W&D): jointly trains a wide linear model and deep neural network, to memorize interactions from high-dimensional sparse features (e.g. bag-of-codes features from EHR) and generalize to unseen feature combinations.
- TabNet^51^: a deep learning model that has performed well across different tabular datasets. It combines the idea of neural network and decision trees by employing a sequential attention mechanism to choose which features to process at each decision step.

We randomly split each cohort data into a training set (75%) and a hold-out test set (25%) while keeping the prevalence of early-onset BD the same before and after the split. Among the training set, 5-fold cross validation was used to optimize LR, NBC and RF. For LGBM, MLP, W&D, and TabNet, we used 20% of the training data as a hold-out validation set. This setup was used for each clinical setting and prediction window. For evaluating model performances, we report area under the receiver operator characteristic curve (AUROC), as well as positive predictive value (PPV) and sensitivity with specificity set to 0.90, 0.95 and 0.99.

In addition, we selected and combined the 20 most important (non-demographic) features (based on global SHAP^52^ values of the validation set, part of the training data) from the best performing models, and trained two “lightweight” models for each cohort. To evaluate the model simplicity vs. accuracy, we selected logistic regression (LR) and Random Forest (RF) for the lightweight models, which we refer to as “LR-S” and “RF-S”. We also repeated this procedure with the top-10 (non-demographic) features and named the resulting models as “LR-XS” and “RF-XS”. The rationale for training these lower-dimensional models was to determine whether simpler models, which might be less computationally burdensome and more portable to other health systems, might perform adequately in comparison to our higher-dimensional machine learning models.

## 3. Results

A total of 300,398, 105,461 and 35,213 patients were included in the general, mental healthcare and mood disorder/ADHD cohorts, respectively, before the landmark sampling. Table 1 shows the demographic breakdown of the three cohorts. In terms of gender distribution, all cohorts had slightly greater representation of females than males (e.g., 58.4% females in the general cohort). All cohorts also had a predominance of self-reported White race (e.g., 66.5% in the general cohort).

**Table 1.** Demographic breakdown of the three study cohorts before landmark sampling with different prediction windows.

| **Setting** | **General** | **Mental healthcare** | **Mood Disorder / ADHD** |
| --- | --- | --- | --- |
| Total *N* | 300,398 | 105,461 | 35,213 |
| Gender: *N* (%) |  |  |  |
| *Female* | 175,427 (58.4) | 59,608 (56.5) | 16,452 (46.7) |
| *Male* | 124,963 (41.6) | 45,851 (43.5) | 18,760 (53.3) |
| *Unknown* | 8 (0.00) | 2 (0.00) | 1 (0.00) |
| Self-reported Race: *N* (%) |  |  |  |
| *Asian* | 12,982 (4.3) | 3,364 (3.2) | 1,095 (3.1) |
| *Black/African American* | 26,086 (8.7) | 9,371 (8.9) | 2,376 (6.8) |
| *White* | 199,777 (66.5) | 68,912 (65.3) | 25,143 (71.4) |
| *Other* | 28,704 (9.6) | 9,928 (9.4) | 3,119 (8.9) |
| *Unknown* | 32,849 (10.9) | 13,886 (13.2) | 3,480 (9.9) |
| Ethnicity: *N* (%) |  |  |  |
| *Hispanic* | 40,317 (13.4) | 16,520 (15.7) | 4,257 (12.1) |
| *Non-Hispanic* | 260,081 (86.6) | 88,941 (84.3) | 30,956 (87.9) |

Table 2 shows the number of patients from the 3 cohorts and 4 different prediction windows after the landmark sampling. The mood disorder/ADHD cohort on average had slightly younger patients than the other two cohorts. Prevalence of early-onset BD was the lowest in the general cohort (ranging from 0.11% for the 6-month prediction to 0.28% for the 3-year prediction windows) and highest in the mood disorder/ADHD cohort (0.40% to 0.89%). The median visit count and CPT code count of the mental health visits and mood disorder/ADHD cohorts were roughly twice as high as those of the general cohort. The median ICD and CPT durations if the mood disorder/ADHD cohort were also twice as long as those in the general cohort.

**Table 2.** Number of patients of the three cohorts after landmark sampling, with four different prediction windows.

| Setting | Pred. win.*<br>(year) | No. of patients | Age, Mean (SD) | Incident BD** | Hospital usage, Median (IQR) |  |  |  |
| --- | --- | --- | --- | --- | --- | --- | --- | --- |
|  |  |  |  |  | Visit count | CPT count | ICD duration<sup>#</sup> | CPT duration<sup>#</sup> |
| General | 0.5 | 288,914 | 18.6 (4.5) | 307 (0.11) | 8 (21) | 21 (44) | 3.44 (6.8) | 3.28 (6.5) |
|  | 1 | 276,810 | 18.3 (4.4) | 432 (0.16) | 8 (15) | 21 (45) | 3.48 (7.0) | 3.31 (6.7) |
|  | 2 | 253,120 | 17.6 (4.1) | 577 (0.23) | 8 (15) | 20 (44) | 3.56 (7.4) | 3.38 (7.1) |
|  | 3 | 232,099 | 17.0 (3.8) | 653 (0.28) | 7 (16) | 19 (45) | 3.65 (7.8) | 3.44 (7.5) |
| Mental Healthcare | 0.5 | 101,798 | 18.8 (4.5) | 306 (0.30) | 16 (30) | 42 (86) | 5.01 (8.3) | 4.83 (8.2) |
|  | 1 | 97,415 | 18.4 (4.3) | 441 (0.45) | 16 (31) | 41 (86) | 5.06 (8.3) | 4.88 (8.2) |
|  | 2 | 88,114 | 17.7 (4.1) | 519 (0.59) | 16 (31) | 41 (86) | 5.27 (8.4) | 5.07 (8.4) |
|  | 3 | 79,507 | 17.0 (3.8) | 575 (0.72) | 17 (33) | 40 (88) | 5.55 (8.6) | 5.32 (8.5) |
| Mood Disorder /ADHD | 0.5 | 34,364 | 17.6 (4.5) | 137 (0.40) | 18 (35) | 36 (87) | 6.58 (9.1) | 6.31 (9.0) |
|  | 1 | 33,326 | 17.3 (4.3) | 193 (0.58) | 18 (36) | 36 (87) | 6.61 (9.1) | 6.34 (9.0) |
|  | 2 | 30,983 | 16.8 (4.0) | 225 (0.73) | 18 (37) | 35 (87) | 6.83 (9.0) | 6.56 (8.9) |
|  | 3 | 28,849 | 16.2 (3.7) | 258 (0.89) | 18 (38) | 34 (87) | 6.93 (9.0) | 6.67 (8.9) |

\*Pred. win.: prediction windows used in each experiment, ranging from 6 months to 3 years.
\*\*Incident BD: the number (%) of patients who had a first documented diagnosis of BD within the prediction window.
<sup>#</sup>ICD and CPT duration are calculated by subtracting the recorded dates of the patients' first ICD and CPT codes from their sampled landmark visit dates.

Overall, of the 7 models, the two tree-based models (RF and LGBM) consistently yielded the best performance. For example, the RF model had an AUROC ranging from 0.76 to 0.88 across different cohorts and prediction windows, compared to 0.64 – 0.77 for LR, 0.65 – 0.81 for NBC, and 0.64 – 0.81 for the TabNet model. At 95% specificity, the tree-based models also outperformed the other models in PPV across different settings, with the highest PPV (4.5%) observed in the mental healthcare cohort using a 3-year prediction window. Given this, we only show the performance comparison between the two tree-based models in Table 3, but results for all 7 models are shown in Supplementary Tables 1–3. The two models performed similarly in the general and mental healthcare cohorts, with RF outperforming LGBM for the mood disorder/ADHD cohort (AUROCs of 0.76 – 0.83 and PPVs of 2% - 4.3% at 95% specificity). Although the general cohort had the lowest PPVs (as expected given their lower base rate of incident BD), those classified as high risk were at least 7 times more likely to have early-onset BD compared to those classified as low risk. Because clinicians may be most concerned about patients with the highest predicted risk, we also evaluated the RF models’ performance in identifying such patients. For example, Supplementary Figure 2 (b) shows the cumulative gain charts for 1 year prediction window. For patients in the top quintile of predicted risk, the model identifies more than 80% of the early-onset BD cases in the general cohort, and more than 60% for the other two cohorts. Similar results were observed with other prediction windows.

**Table 3.** Model performance comparison between Random Forest (RF) and LightGBM (LGBM), for four prediction windows (Pred. win.) and three cohorts: (a) general patients; (b) mental healthcare; (c) mood disorder/ADHD. Sensitivity, positive predictive value (PPV) and relative risk (RR) are reported with 95% specificity.

| (a) General cohort |  |  |  |  |  |  |
| --- | --- | --- | --- | --- | --- | --- |
| Model | Pred. win. | AUROC | Sensitivity | PPV | RR | Prevalence |
| RF | 6 months | 0.873 | 0.532 | 1.0% | 9.80 | 0.11% |
| LGBM | 6 months | 0.845 | 0.506 | 1.0% | 9.71 | 0.11% |
| RF | 1 year | 0.880 | 0.481 | 1.5% | 9.48 | 0.16% |
| LGBM | 1 year | 0.886 | 0.509 | 1.4% | 9.28 | 0.16% |
| RF | 2 years | 0.833 | 0.472 | 2.1% | 9.14 | 0.23% |
| LGBM | 2 years | 0.802 | 0.417 | 1.8% | 7.98 | 0.23% |
| RF | 3 years | 0.801 | 0.411 | 2.2% | 7.95 | 0.28% |
| LGBM | 3 years | 0.793 | 0.411 | 2.2% | 7.99 | 0.28% |
| (b) Mental healthcare cohort |  |  |  |  |  |  |
| Model | Pred. win. | AUROC | Sensitivity | PPV | RR | Prevalence |
| RF | 6 months | 0.778 | 0.247 | 1.4% | 4.65 | 0.30% |
| LGBM | 6 months | 0.758 | 0.273 | 1.6% | 5.35 | 0.30% |
| RF | 1 year | 0.796 | 0.255 | 2.2% | 4.79 | 0.45% |
| LGBM | 1 year | 0.785 | 0.309 | 2.7% | 5.93 | 0.45% |
| RF | 2 years | 0.762 | 0.231 | 2.6% | 4.33 | 0.59% |
| LGBM | 2 years | 0.750 | 0.231 | 2.5% | 4.21 | 0.59% |
| RF | 3 years | 0.808 | 0.333 | 4.5% | 6.16 | 0.72% |
| LGBM | 3 years | 0.790 | 0.319 | 4.3% | 5.99 | 0.72% |
| (c) Mood disorder/ADHD cohort |  |  |  |  |  |  |
| Model | Pred. win. | AUROC | Sensitivity | PPV | RR | Prevalence |
| RF | 6 months | 0.791 | 0.324 | 2.0% | 5.13 | 0.40% |
| LGBM | 6 months | 0.774 | 0.294 | 2.3% | 5.69 | 0.40% |
| RF | 1 year | 0.827 | 0.354 | 3.8% | 6.61 | 0.58% |
| LGBM | 1 year | 0.763 | 0.333 | 3.4% | 5.90 | 0.58% |
| RF | 2 years | 0.800 | 0.321 | 4.3% | 5.89 | 0.73% |
| LGBM | 2 years | 0.741 | 0.286 | 3.7% | 5.03 | 0.73% |
| RF | 3 years | 0.757 | 0.215 | 3.6% | 4.00 | 0.89% |
| LGBM | 3 years | 0.747 | 0.154 | 2.6% | 2.95 | 0.89% |

To illustrate feature importance, Figure 3 shows the SHAP summary plots from the RF models and the 2-year prediction window generated by taking the mean absolute SHAP value of the test data. (SHAP plots for the other prediction windows are shown in Supplementary Figures 7–9). Having higher numbers of phecode counts for depression and mood disorders (excluding BD), and fewer NLP instances of “disruptive, impulse control and conduct disorders” (DICD) appeared to be the most important features across different cohorts for predicting early onset BD (using the 2-year prediction window). The importance of other features varied across cohorts (Figure 3).

**Figure 3:**
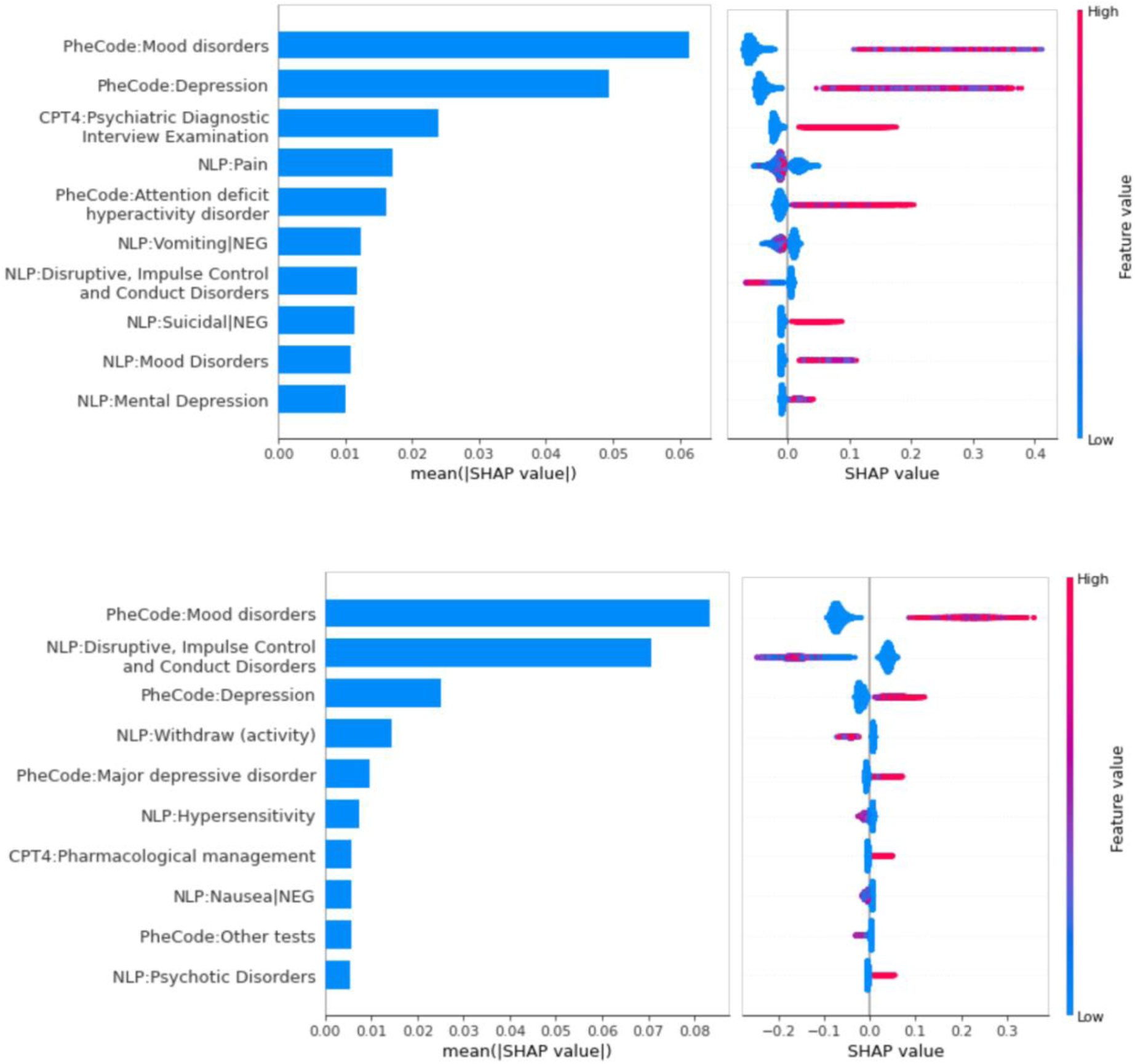
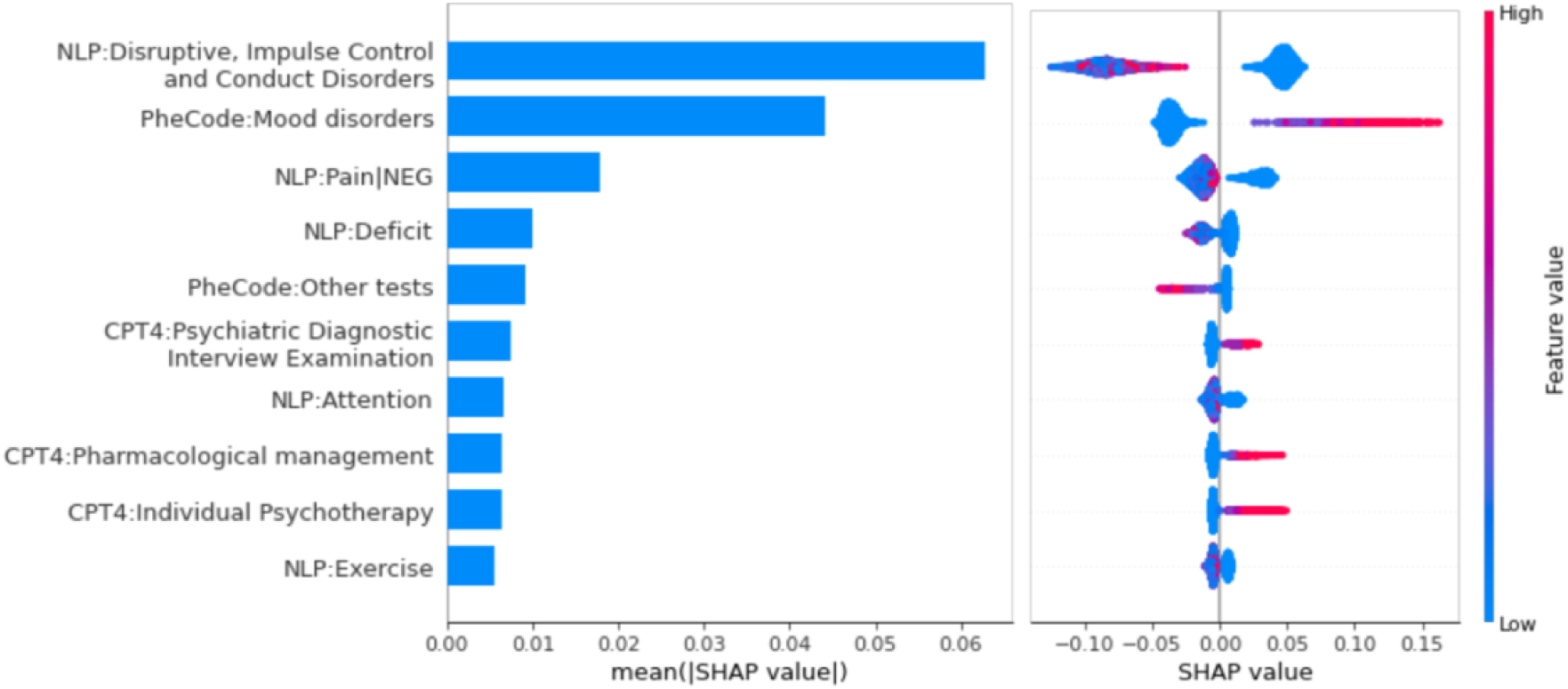
SHAP summary plots of the top-10 features for the General cohort (a), mental healthcare (b), and mood disorder/ADHD cohort (c) for the 2-year prediction window. The global importance of each feature is determined by the mean absolute Shapley value for that feature over all the test samples. The beeswarm plot on the right shows the relationship between the value of a feature and the impact on the prediction. NEG indicates that the NLP feature was negated.

In exploratory analyses (Supplementary Figures 3–6), we examined model performance stratified by demographic variables for the RF model with a 2-year prediction window: age (10-17 years vs. 18-24 years), number of visits, gender, and self-reported race (White vs. non-White). With respect to age group, model performance (by AUC, PPV and sensitivity, RR) was greater for younger patients (age 10-18) across the three cohorts. Performance also tended to be greater for patients with more than 10 hospital visits, likely due to the greater availability of longitudinal features. Overall, despite some variations, model performance was generally consistent when stratified by gender and race groups, except for the mood disorder/ADHD cohort where performance point estimates were higher for female and non-White groups.

For the S/XS models, we selected and combined the top-20/10 non-demographic features from the two best performing models, i.e., RF and LGBM, resulting in 58 S/48 XS features for the general cohort, 62/47 features for the mental healthcare cohort, and 70/51 features for the mood disorder/ADHD cohort, respectively, for the 2-year prediction task. Figure 4 shows the model performance for the lightweight models, LR-XS, LR-S, RF-XS and RF-S, as well as their full model (LR-full and RF-full) counterparts. The RF-XS model performed very similarly to the full model by different metrics. Of note, the LR-XS and LR-S models outperformed the full LR model, possibly due to a large number of irrelevant or redundant features in the LR-full model. Results for the other prediction windows are shown in Supplementary Figures 10–12.

**Figure 4:**
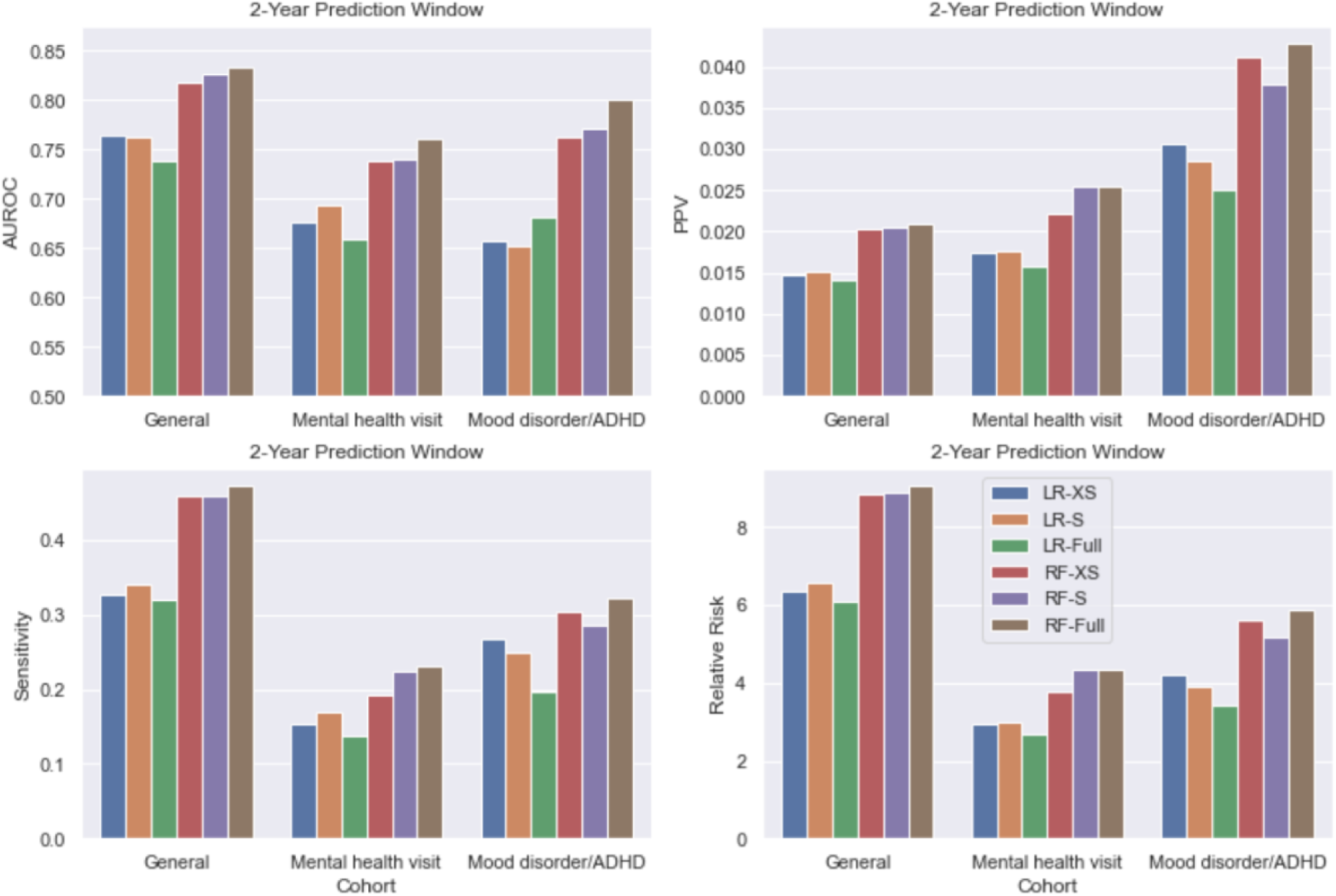
Model performance comparison between the original RF model using the full list of features and the light-weight models, with 2-year prediction window.

## 4. Discussion

Earlier age of onset of BD has been associated with a more severe course, heightened suicidal risk, and poorer functional outcomes^53^, and early intervention has been reported to improve prognosis^54,55^. However, there are currently no well-established tools to stratify risk of BD onset. Moreover, when BD symptoms do emerge, they might initially present as depressive episodes.

Particularly in younger patients, ADHD can complicate differential diagnosis due to clinical features (e.g. distractibility, hyperactivity) that might overlap with prodromal bipolar symptoms. Given these complexities, early detection and intervention for early-onset BD remain clinical challenges.

Prior attempts to identify youth at risk for BD have typically relied on structured assessments or deep phenotyping that may not be commonly used in clinical practice or may be difficult to scale^13,14^. The availability of large-scale, real-world healthcare data and progress in machine learning has provided an opportunity to develop accurate, scalable tools for risk stratification and screening. A recent study used such methods to develop EHR-based algorithms for the prediction of BD but those analyses were restricted to adult patients^56^. In this study, we developed various machine learning models to identify youth at risk of BD for three clinical use cases: a general cohort of all youth in a health system, youth with a history of mental healthcare, and youth with prior diagnosis of a (non-BD) mood disorder or ADHD. We utilized large-scale EHR data including features extracted from unstructured clinical notes, and systematically examined their performance for the prediction of early-onset BD (documented diagnosis before age 25). We adopted a “landmark model” framework^24–27^ that addresses temporal bias in case-control sampling^21^, enabling us to conduct predictive modeling in a manner closely aligned with real-world scenarios where a clinician would want to assess prospective risk. Both Random Forest (RF) and LightGBM (LGBM) models achieved good discrimination performance across the 3 patient cohorts and 4 prediction windows ranging from 6 months to 3 years. In addition, “lightweight” RF-S/-XS models with a greatly reduced set of features also achieved comparable performance, demonstrating that computational efficiency can be attained without significant compromise on model accuracy.

Notably, the two tree-based models, RF and LGBM, achieving the best model discrimination (e.g., AUC=0.74-0.89) across all cohorts compared to other benchmarked models including three neural network-based models. However, we also observed trade-offs of discrimination and precision (PPV) based on cohort differences in the base-rate of BD. As shown in Table 3, the highest AUCs (0.79-0.89) and sensitivity (0.41-0.51 at 95% specificity) were observed for the general cohort, in which patients classified as high-risk had a 7- to 9-fold increased risk (at 95% specificity) of early-onset BD across different prediction windows. On the other hand, the general cohort had the lowest PPVs, ranging from 1% to 2.2%. In contrast, higher PPVs but lower sensitivity and AUCs were observed for the mental health (AUC: 0.75-0.81, sensitivity: 0.23-0.33, PPV: 1.4%-4.5%) and mood disorder/ADHD (AUC: 0.74-0.83, sensitivity: 0.15-0.35, PPV: 2.0%-4.3%). This trade-off in performance metrics is due to the variable incidence of early-onset BD in the three cohorts. Notably, we found that “lightweight” versions of the original RF models (based on substantially reduced feature sets) performed as well or even better than the full feature models. In settings where model training efficiency and portability are prioritized, the lightweight models offer feasibility advantages and may also facilitate more frequent updating of model weights.

Several caveats are worth noting regarding the interpretation of the results presented here. First, while feature importance (by mean SHAP values) was largely consistent across prediction windows within the cohorts, it varied by cohort in some respects. This is likely due to variation in the network of feature correlations that distinguish the general pediatric population from patients with prior psychiatric care or illness. It is also important to note that our prediction models are not causal models, and feature importance does not necessarily reflect causal effects of specific features. Thus, intervening on highly predictive features may or may not reduce risk. Second, the relatively modest PPVs observed with even our best performing (RF) models reflect the relatively low base rate of incident diagnosis of BD (11/1000 – 89/1000). Given this, we view the use case for our models as a screening tool to alert clinicians to the need for further evaluation and monitoring for undiagnosed or incipient BD. It would be inappropriate to base clinical decisions solely on model readouts. Third, because our predictions target was a documented diagnosis of BD, a positive modeled outcome can convey two potential meanings: the emergence of BD at a future time point (predicted incident BD) or the detection of undiagnosed BD. Distinguishing these two scenarios for a given patient may not be possible, although a high predicted probability in a child or adolescent patient with no mental health symptoms would more likely represent a future risk of disorder.

There are several other limitations of the presented study: (1) because patients may receive care outside the healthcare system, it is possible that an initial BD diagnosis could be missed. If this were common, it would presumably limit the performance of our models; (2) our models were trained and validated in a single healthcare system, and performance in other systems may vary depending on variation in patient characteristics and documentation practices. Nevertheless, we note that the MGB system comprises more than 8 hospitals and multiple community health centers with heterogeneous catchment areas and clinical practices, supporting some degree of generalizability; (3) some seemingly irrelevant NLP features were selected as important features by SHAP, e.g., NLP CUIs for Hawaii|NEG (negated mention of Hawaii) and International System of Units|NEG (negated mention of international system of units), as seen in Supplementary Figures 7–9. Presumably, these features are not themselves risk factors for BD but rather are “tagging” true but unknown causal factors.

Future work will be needed to evaluate the portability of our approach to other healthcare systems. In addition, there may be opportunities to improve model performance by incorporating additional data types. For example, a recent study^14^ suggests that features derived from parent-reported questionnaires (e.g. the Child behavior checklist (CBCL)) can be used to predict the future development of BD in children and adolescents with emergent psychopathology. Another recent study of 1091 Brazilian youth^57^ applied machine learning to a range sociodemographic and questionnaire data to predict incident BD (n = 49 cases) at 5-year follow-up. It will also be important to explore considerations regarding the possible implementation of our risk models. For example, the net benefit of our such models may vary by setting and whether the goal is to minimize false positives (which might favor our mental healthcare or mood/ADHD models) or false negatives (in which case the greater sensitivity of our general cohort model might be preferred).

In conclusion, we developed and validated risk prediction models using large-scale, real-world EHR data including clinical notes, for predicting bipolar disorder risk among three youth cohorts. Using a prospective landmark modeling approach that mirrors the clinical situation in which future outcomes are not known, we found that tree-based models achieved good predictive performance across different clinical settings. Our approach offers a scalable, accurate method for identifying youth at risk for BD that could inform clinical decision making and facilitate early intervention.

## Data availability

Protected Health Information restrictions apply to the availability of the clinical data here, which were used under IRB approval for use only in the current study. As a result, this dataset is not publicly available.

## Ethics and consent statement

The authors assert that all procedures contributing to this work comply with the ethical standards of the relevant national and institutional committees on human experimentation and with the Helsinki Declaration of 1975, as revised in 2008. The study protocol was approved by the Institutional Review Board of Mass General Brigham (MGB) with a waiver of consent for the analysis of electronic health record data.

## Acknowledgments

This work was supported in part by a gift from the Ryan Licht Sang Bipolar Foundation and NIMH R01MH118233 (JWS).

