## Supplementary Information for "Machine Learning Models for the Prediction of Early-Onset Bipolar Using Electronic Health Records"

**Appendix A1**: ICCBD Coded-Broad definition for early onset bipolar disorder.

**Appendix A2**: Definition of mental health visit.

**Appendix A3**: Definition of mood disorder and ADHD.

**Appendix B**: NLP feature extraction.

**Appendix C**: Model implementation details.

**Supplementary Table 1**: Full model performance comparison among 7 models, for the general patient cohort.

**Supplementary Table 2**: Full model performance comparison among 7 models, for the mental healthcare cohort.

**Supplementary Table 3**: Full model performance comparison among 7 models, for the mood disorder/ADHD cohort.

**Supplementary Figure 1**: Full data processing and cohort building chart flow.

**Supplementary Figure 2**: Cumulative gain charts.

**Supplementary Figure 3**: Model metrics of Random Forest-full (RF-full) using 2-year prediction window, stratified by patients’ age at the landmark visit.

**Supplementary Figure 4**: Model metrics of RF-full using 2-year prediction window, stratified by number of visits each patient had before their “landmark visits”.

**Supplementary Figure 5**: Model metrics of RF-full using 2-year prediction window, stratified by gender (i.e., male vs. female).

**Supplementary Figure 6**: Model metrics of RF-full using 2-year prediction window, stratified by race (i.e., white vs. non-white).

**Supplementary Figure 7**: SHAP summary plots of the top-10 features with 6 months prediction window, for **(a)** the general patient cohorts, **(b)** the mental healthcare cohort, and **(c)** the mood disorder/ADHD cohort.

**Supplementary Figure 8**: SHAP summary plots of the top-10 features with 1-year prediction window, for **(a)** the general patient cohorts, **(b)** the mental healthcare cohort, and **(c)** the mood disorder/ADHD cohort.

**Supplementary Figure 9**: SHAP summary plots of the top-10 features with 3-year prediction window, for **(a)** the general patient cohorts, **(b)** the mental healthcare cohort, and **(c)** the mood disorder/ADHD cohort.

**Supplementary Figure 10**: Model performance comparison between the original RF model and the light-weight models, with 6 months prediction window.

**Supplementary Figure 11**: Model performance comparison between the original RF model and the light-weight models, with 1-year prediction window.

**Supplementary Figure 12**: Model performance comparison between the original RF model and the light-weight models, with 3-year prediction window.

### Appendix A: Various definitions

#### A.1 ICCBD Coded-Broad definition for early onset BD

ICCBD Coded-Broad definition for early onset BD has a data floor – each patient must have at least 3 inpatient, outpatient, or emergency visits over a minimum 180-day window from the first to the last visit.

- For those who later developed early-onset BD (cases): the “last visit” is their first BD diagnosis (i.e., index visit) before age 25.
- For the others: the “last visit” is their last diagnosis date in the EHR before age 25.

The original ICCBD Coded-Broad case/control definitions are:

a. Case definition:
   i. History of:
      1. >=2 BD diagnoses (at separate visits >1 month apart).
      2. No major depressive disorder (MDD), schizophrenia (SCZ), schizoaffective disorder (SZA), or organic affective syndrome (OAS) diagnoses unless 2 most recent diagnoses are BD; or number of MDD, SCZ, SZA, or OAS diagnoses is greater (>50%) than number of BD diagnoses.
      3. >=2 BD medication instances (lithium, valproic acid, risperidone, olanzapine, quetiapine, ziprasidone, aripiprazole, carbamazepine, or lamotrigine) within 1 year of a BD diagnosis.
b. Control definition:
   i. Absence of case status.
   ii. Meets the minimum data floor as above.

#### A.2 Definition of mental health visit

Mental health visits are defined using the ICD codes below:

- ICD-9 290-316 codes
- ICD-10 F codes

#### A.3 Definition of mood disorder and ADHD

Mood disorder excluding bipolar disorder is defined using the ICD codes below:

- ICD-9 262.2
- ICD-9 262.20
- ICD-9 262.21
- ICD-9 262.22
- ICD-9 262.23
- ICD-9 262.24
- ICD-9 262.25
- ICD-9 262.26
- ICD-9 262.3
- ICD-9 262.30
- ICD-9 262.31
- ICD-9 262.32
- ICD-9 262.33
- ICD-9 262.34
- ICD-9 262.35
- ICD-9 262.36
- ICD-9 311
- ICD-9 300.4
- ICD-9 296.9
- ICD-9 296.90
- ICD-9 296.99
- ICD-10 F32.0
- ICD-10 F32.1
- ICD-10 F32.2
- ICD-10 F32.3
- ICD-10 F32.4
- ICD-10 F32.5
- ICD-10 F32.8
- ICD-10 F32.81
- ICD-10 F32.9
- ICD-10 F32.A
- ICD-10 F33.0
- ICD-10 F33.1
- ICD-10 F33.2
- ICD-10 F33.3
- ICD-10 F33.4
- ICD-10 F33.40
- ICD-10 F33.41
- ICD-10 F33.42
- ICD-10 F33.8
- ICD-10 F33.9
- ICD-10 F34.1
- ICD-10 F34.8
- ICD-10 F34.81
- ICD-10 F34.89
- ICD-10 F34.9

ADHD is defined using the ICD codes below:

- ICD-9 314
- ICD-9 314.0
- ICD-9 314.00
- ICD-9 314.01
- ICD-9 314.1
- ICD-9 314.2
- ICD-9 314.8
- ICD-9 314.9
- ICD-10 F90.0
- ICD-10 F90.1
- ICD-10 F90.2
- ICD-10 F90.8
- ICD-10 F90.9

### Appendix B: NLP feature extraction

The rule-based system is based on a custom lexicon of mental health related concepts using a variety of approaches: (1) selecting relevant semantic types from the Unified Medical Language System (UMLS)[^11^](https://sciwheel.com/work/citation?ids=267431&pre=&suf=&sa=0&dbf=0); (2) mapping of Diagnostic and Statistical Manual of Mental Disorders (DSM) symptoms and concepts from structured instruments[^39^](https://sciwheel.com/work/citation?ids=14939058&pre=&suf=&sa=0&dbf=0); (3) applying automated feature extraction from public sources including Wikipedia and MedScape; (4) incorporating Research Domain Criteria (RDoC) domain matrix terms[^40^](https://sciwheel.com/work/citation?ids=4913386&pre=&suf=&sa=0&dbf=0); (5) selecting predictive features from coded suicide attempt prediction models[^15^](https://sciwheel.com/work/citation?ids=3155377&pre=&suf=&sa=0&dbf=0); (6) manual annotation terms by clinical reviewers. Using this lexicon, we ran the HiTex[^41^](https://sciwheel.com/work/citation?ids=3718385&pre=&suf=&sa=0&dbf=0) NLP named entity extraction (NER) pipeline to identify concepts in over 120 million clinical notes and applied the ConText algorithm[^42^](https://sciwheel.com/work/citation?ids=14939054&pre=&suf=&sa=0&dbf=0) for negation and family history recognition.

### Appendix C: Model implementation details

We used the *scikit-learn*[^43^](https://sciwheel.com/work/citation?ids=14871511&pre=&suf=&sa=0&dbf=0) implementation of LR and NBC, the Python library *imblearn*[^44^](https://sciwheel.com/work/citation?ids=14871609&pre=&suf=&sa=0&dbf=0) for a balanced random forest classifier, and the original Python implementation *lightgbm* for LGBM. We implemented MLP and W&D using the Python deep learning library, *keras*, and adopted a *PyTorch* implementation of TabNet (<https://github.com/dreamquark-ai/tabnet>). Hyperparameter tuning was performed using *Optuna*[^45^](https://sciwheel.com/work/citation?ids=7850897&pre=&suf=&sa=0&dbf=0) with 100 trials for each model.

All models were trained and tested on a dedicated Windows 10 server running with 8-core Intel Xeon W-2145 CPU @ 3.70GHz and 16 logical processors, 512 GB of memory (RAM) and a RTX 2080 Ti.

### Supplementary Materials

##### **Supplementary Table 1.** Full model performance comparison among 7 models, for the general patient cohort.

| 6 months prediction window (prevalence = 0.11%) | | | | | | | |
| --- | --- | --- | --- | --- | --- | --- | --- |
| Model | 80% spec. | | 90% spec. | | 95% spec. | | AUROC |
| LogReg | PPV | 0.003 | PPV | 0.005 | PPV | 0.006 | 0.768 |
|  | Sen. | 0.597 | Sen. | 0.494 | Sen. | 0.299 |  |
| NBC | PPV | 0.003 | PPV | 0.005 | PPV | 0.009 | 0.812 |
|  | Sen. | 0.714 | Sen. | 0.519 | Sen. | 0.429 |  |
| RF | PPV | **0.004** | PPV | **0.007** | PPV | **0.010** | **0.873** |
|  | Sen. | **0.805** | Sen. | **0.688** | Sen. | **0.532** |  |
| LGBM | PPV | 0.004 | PPV | 0.006 | PPV | **0.010** | 0.845 |
|  | Sen. | 0.779 | Sen. | 0.597 | Sen. | 0.506 |  |
| MLP | PPV | 0.003 | PPV | 0.005 | PPV | 0.008 | 0.775 |
|  | Sen. | 0.610 | Sen. | 0.506 | Sen. | 0.364 |  |
| W&D | PPV | 0.003 | PPV | 0.005 | PPV | 0.006 | 0.753 |
|  | Sen. | 0.571 | Sen. | 0.468 | Sen. | 0.286 |  |
| TabNet | PPV | 0.004 | PPV | 0.006 | PPV | 0.009 | 0.806 |
|  | Sen. | 0.753 | Sen. | 0.571 | Sen. | 0.442 |  |
| 1 year prediction window (prevalence = 0.16%) | | | | | | | |
| Model | 80% spec. | | 90% spec. | | 95% spec. | | AUROC |
| LogReg | PPV | 0.004 | PPV | 0.006 | PPV | 0.010 | 0.767 |
|  | Sen. | 0.556 | Sen. | 0.435 | Sen. | 0.398 |  |
| NBC | PPV | 0.005 | PPV | 0.008 | PPV | 0.010 | 0.799 |
|  | Sen. | 0.630 | Sen. | 0.519 | Sen. | 0.343 |  |
| RF | PPV | **0.006** | PPV | **0.010** | PPV | **0.015** | 0.880 |
|  | Sen. | 0.833 | Sen. | 0.676 | Sen. | 0.481 |  |
| LGBM | PPV | **0.006** | PPV | **0.010** | PPV | 0.014 | **0.886** |
|  | Sen. | **0.852** | Sen. | **0.694** | Sen. | **0.509** |  |
| MLP | PPV | 0.005 | PPV | 0.006 | PPV | 0.009 | 0.762 |
|  | Sen. | 0.593 | Sen. | 0.417 | Sen. | 0.287 |  |
| W&D | PPV | 0.004 | PPV | 0.006 | PPV | 0.009 | 0.723 |
|  | Sen. | 0.556 | Sen. | 0.398 | Sen. | 0.306 |  |
| TabNet | PPV | 0.006 | PPV | 0.007 | PPV | 0.013 | 0.807 |
|  | Sen. | 0.713 | Sen. | 0.500 | Sen. | 0.435 |  |
| 2 years prediction window (prevalence = 0.23%) | | | | | | | |
| Model | 80% spec. | | 90% spec. | | 95% spec. | | AUROC |
| LogReg | PPV | 0.006 | PPV | 0.010 | PPV | 0.014 | 0.738 |
|  | Sen. | 0.569 | Sen. | 0.451 | Sen. | 0.257 |  |
| NBC | PPV | 0.006 | PPV | 0.010 | PPV | 0.011 | 0.717 |
|  | Sen. | 0.549 | Sen. | 0.451 | Sen. | 0.257 |  |
| RF | PPV | **0.008** | PPV | **0.014** | PPV | **0.021** | **0.833** |
|  | Sen. | **0.771** | Sen. | **0.653** | Sen. | **0.472** |  |
| LGBM | PPV | **0.008** | PPV | **0.014** | PPV | 0.018 | 0.802 |
|  | Sen. | 0.694 | Sen. | 0.618 | Sen. | 0.417 |  |
| MLP | PPV | 0.007 | PPV | 0.009 | PPV | 0.012 | 0.751 |
|  | Sen. | 0.583 | Sen. | 0.444 | Sen. | 0.278 |  |
| W&D | PPV | 0.006 | PPV | 0.009 | PPV | 0.012 | 0.730 |
|  | Sen. | 0.549 | Sen. | 0.403 | Sen. | 0.278 |  |
| TabNet | PPV | 0.007 | PPV | 0.011 | PPV | 0.014 | 0.765 |
|  | Sen. | 0.667 | Sen. | 0.500 | Sen. | 0.333 |  |
| 3 years prediction window (prevalence = 0.28%) | | | | | | | |
| Model | 80% spec. | | 90% spec. | | 95% spec. | | AUROC |
| LogReg | PPV | 0.008 | PPV | 0.011 | PPV | 0.017 | 0.741 |
|  | Sen. | 0.564 | Sen. | 0.411 | Sen. | 0.307 |  |
| NBC | PPV | 0.008 | PPV | 0.010 | PPV | 0.013 | 0.704 |
|  | Sen. | 0.540 | Sen. | 0.380 | Sen. | 0.233 |  |
| RF | PPV | **0.009** | PPV | **0.015** | PPV | **0.022** | **0.801** |
|  | Sen. | **0.663** | Sen. | **0.546** | Sen. | **0.411** |  |
| LGBM | PPV | **0.009** | PPV | **0.015** | PPV | **0.022** | 0.793 |
|  | Sen. | 0.650 | Sen. | 0.534 | Sen. | **0.411** |  |
| MLP | PPV | 0.008 | PPV | 0.011 | PPV | 0.014 | 0.711 |
|  | Sen. | 0.540 | Sen. | 0.387 | Sen. | 0.252 |  |
| W&D | PPV | 0.007 | PPV | 0.009 | PPV | 0.013 | 0.701 |
|  | Sen. | 0.521 | Sen. | 0.344 | Sen. | 0.245 |  |
| TabNet | PPV | 0.008 | PPV | 0.012 | PPV | 0.019 | 0.738 |
|  | Sen. | 0.601 | Sen. | 0.454 | Sen. | 0.374 |  |

##### **Supplementary Table 2.** Full model performance comparison among 7 models, for the mental healthcare cohort.

| 6 months prediction window (prevalence = 0.40%) | | | | | | | |
| --- | --- | --- | --- | --- | --- | --- | --- |
| Model | 80% spec. | | 90% spec. | | 95% spec. | | AUROC |
| LogReg | PPV | 0.005 | PPV | 0.006 | PPV | 0.007 | 0.652 |
|  | Sen. | 0.377 | Sen. | 0.195 | Sen. | 0.143 |  |
| NBC | PPV | 0.006 | PPV | 0.006 | PPV | 0.007 | 0.659 |
|  | Sen. | 0.377 | Sen. | 0.221 | Sen. | 0.117 |  |
| RF | PPV | **0.009** | PPV | **0.012** | PPV | 0.014 | **0.777** |
|  | Sen. | **0.610** | Sen. | **0.429** | Sen. | 0.247 |  |
| LGBM | PPV | 0.008 | PPV | 0.011 | PPV | **0.016** | 0.758 |
|  | Sen. | 0.571 | Sen. | 0.377 | Sen. | **0.273** |  |
| MLP | PPV | 0.005 | PPV | 0.006 | PPV | 0.006 | 0.624 |
|  | Sen. | 0.364 | Sen. | 0.208 | Sen. | 0.091 |  |
| W&D | PPV | 0.005 | PPV | 0.005 | PPV | 0.005 | 0.637 |
|  | Sen. | 0.364 | Sen. | 0.156 | Sen. | 0.078 |  |
| TabNet | PPV | 0.007 | PPV | 0.007 | PPV | 0.007 | 0.720 |
|  | Sen. | 0.468 | Sen. | 0.247 | Sen. | 0.117 |  |
| 1 year prediction window (prevalence = 0.58%) | | | | | | | |
| Model | 80% spec. | | 90% spec. | | 95% spec. | | AUROC |
| LogReg | PPV | 0.011 | PPV | 0.015 | PPV | 0.021 | 0.716 |
|  | Sen. | 0.500 | Sen. | 0.364 | Sen. | 0.245 |  |
| NBC | PPV | 0.011 | PPV | 0.015 | PPV | 0.018 | 0.726 |
|  | Sen. | 0.509 | Sen. | 0.345 | Sen. | 0.209 |  |
| RF | PPV | **0.015** | PPV | 0.019 | PPV | 0.022 | **0.796** |
|  | Sen. | **0.709** | Sen. | 0.445 | Sen. | 0.255 |  |
| LGBM | PPV | 0.014 | PPV | **0.021** | PPV | **0.027** | 0.785 |
|  | Sen. | 0.645 | Sen. | **0.482** | Sen. | **0.309** |  |
| MLP | PPV | 0.010 | PPV | 0.012 | PPV | 0.018 | 0.692 |
|  | Sen. | 0.473 | Sen. | 0.300 | Sen. | 0.209 |  |
| W&D | PPV | 0.011 | PPV | 0.012 | PPV | 0.015 | 0.692 |
|  | Sen. | 0.473 | Sen. | 0.282 | Sen. | 0.164 |  |
| TabNet | PPV | 0.011 | PPV | 0.014 | PPV | 0.019 | 0.710 |
|  | Sen. | 0.500 | Sen. | 0.327 | Sen. | 0.218 |  |
| 2 years prediction window (prevalence = 0.73%) | | | | | | | |
| Model | 80% spec. | | 90% spec. | | 95% spec. | | AUROC |
| LogReg | PPV | 0.011 | PPV | 0.015 | PPV | 0.016 | 0.660 |
|  | Sen. | 0.400 | Sen. | 0.254 | Sen. | 0.138 |  |
| NBC | PPV | 0.010 | PPV | 0.012 | PPV | 0.009 | 0.647 |
|  | Sen. | 0.354 | Sen. | 0.231 | Sen. | 0.077 |  |
| RF | PPV | **0.016** | PPV | 0.018 | PPV | **0.026** | **0.761** |
|  | Sen. | **0.546** | Sen. | 0.308 | Sen. | **0.231** |  |
| LGBM | PPV | **0.016** | PPV | **0.021** | PPV | 0.025 | 0.750 |
|  | Sen. | **0.546** | Sen. | **0.377** | Sen. | **0.231** |  |
| MLP | PPV | 0.009 | PPV | 0.011 | PPV | 0.016 | 0.648 |
|  | Sen. | 0.331 | Sen. | 0.200 | Sen. | 0.146 |  |
| W&D | PPV | 0.010 | PPV | 0.010 | PPV | 0.012 | 0.626 |
|  | Sen. | 0.338 | Sen. | 0.177 | Sen. | 0.100 |  |
| TabNet | PPV | 0.013 | PPV | 0.017 | PPV | 0.021 | 0.670 |
|  | Sen. | 0.446 | Sen. | 0.292 | Sen. | 0.192 |  |
| 3 years prediction window (prevalence = 0.89%) | | | | | | | |
| Model | 80% spec. | | 90% spec. | | 95% spec. | | AUROC |
| LogReg | PPV | 0.019 | PPV | 0.024 | PPV | 0.030 | 0.723 |
|  | Sen. | 0.528 | Sen. | 0.347 | Sen. | 0.222 |  |
| NBC | PPV | 0.016 | PPV | 0.018 | PPV | 0.024 | 0.687 |
|  | Sen. | 0.458 | Sen. | 0.257 | Sen. | 0.174 |  |
| RF | PPV | 0.021 | PPV | **0.032** | PPV | **0.045** | **0.808** |
|  | Sen. | 0.597 | Sen. | **0.458** | Sen. | **0.333** |  |
| LGBM | PPV | **0.022** | PPV | 0.030 | PPV | 0.043 | 0.790 |
|  | Sen. | **0.611** | Sen. | 0.438 | Sen. | 0.319 |  |
| MLP | PPV | 0.016 | PPV | 0.020 | PPV | 0.023 | 0.696 |
|  | Sen. | 0.458 | Sen. | 0.319 | Sen. | 0.167 |  |
| W&D | PPV | 0.017 | PPV | 0.018 | PPV | 0.021 | 0.700 |
|  | Sen. | 0.472 | Sen. | 0.264 | Sen. | 0.153 |  |
| TabNet | PPV | 0.019 | PPV | 0.025 | PPV | 0.031 | 0.758 |
|  | Sen. | 0.549 | Sen. | 0.347 | Sen. | 0.222 |  |

##### **Supplementary Table 3.** Full model performance comparison among 7 models, for the mood disorder/ADHD cohort.

| 6 months prediction window (prevalence = 0.40%) | | | | | | | |
| --- | --- | --- | --- | --- | --- | --- | --- |
| Model | 80% spec. | | 90% spec. | | 95% spec. | | AUROC |
| LogReg | PPV | 0.007 | PPV | 0.009 | PPV | 0.009 | 0.635 |
|  | Sen. | 0.412 | Sen. | 0.235 | Sen. | 0.118 |  |
| NBC | PPV | 0.007 | PPV | 0.010 | PPV | 0.008 | 0.654 |
|  | Sen. | 0.353 | Sen. | 0.265 | Sen. | 0.118 |  |
| RF | PPV | 0.012 | PPV | 0.013 | PPV | 0.021 | **0.791** |
|  | Sen. | 0.676 | Sen. | 0.382 | Sen. | **0.324** |  |
| LGBM | PPV | **0.014** | PPV | **0.016** | PPV | **0.023** | 0.774 |
|  | Sen. | **0.706** | Sen. | **0.412** | Sen. | 0.294 |  |
| MLP | PPV | 0.005 | PPV | 0.007 | PPV | 0.010 | 0.605 |
|  | Sen. | 0.294 | Sen. | 0.235 | Sen. | 0.176 |  |
| W&D | PPV | 0.005 | PPV | 0.005 | PPV | 0.007 | 0.603 |
|  | Sen. | 0.265 | Sen. | 0.118 | Sen. | 0.088 |  |
| TabNet | PPV | 0.010 | PPV | 0.010 | PPV | 0.014 | 0.675 |
|  | Sen. | 0.529 | Sen. | 0.353 | Sen. | 0.206 |  |
| 1 year prediction window (prevalence = 0.58%) | | | | | | | |
| Model | 80% spec. | | 90% spec. | | 95% spec. | | AUROC |
| LogReg | PPV | 0.012 | PPV | 0.016 | PPV | 0.018 | 0.667 |
|  | Sen. | 0.458 | Sen. | 0.292 | Sen. | 0.188 |  |
| NBC | PPV | 0.013 | PPV | 0.016 | PPV | 0.012 | 0.650 |
|  | Sen. | 0.458 | Sen. | 0.292 | Sen. | 0.188 |  |
| RF | PPV | **0.018** | PPV | **0.025** | PPV | **0.038** | **0.827** |
|  | Sen. | **0.667** | Sen. | **0.542** | Sen. | **0.354** |  |
| LGBM | PPV | 0.015 | PPV | 0.021 | PPV | 0.034 | 0.763 |
|  | Sen. | 0.563 | Sen. | 0.417 | Sen. | 0.333 |  |
| MLP | PPV | 0.011 | PPV | 0.014 | PPV | 0.034 | 0.655 |
|  | Sen. | 0.417 | Sen. | 0.292 | Sen. | 0.146 |  |
| W&D | PPV | 0.013 | PPV | 0.017 | PPV | 0.022 | 0.668 |
|  | Sen. | 0.458 | Sen. | 0.333 | Sen. | 0.208 |  |
| TabNet | PPV | 0.015 | PPV | 0.016 | PPV | 0.017 | 0.727 |
|  | Sen. | 0.542 | Sen. | 0.292 | Sen. | 0.146 |  |
| 2 years prediction window (prevalence = 0.73%) | | | | | | | |
| Model | 80% spec. | | 90% spec. | | 95% spec. | | AUROC |
| LogReg | PPV | 0.016 | PPV | 0.020 | PPV | 0.025 | 0.681 |
|  | Sen. | 0.482 | Sen. | 0.304 | Sen. | 0.196 |  |
| NBC | PPV | 0.018 | PPV | 0.023 | PPV | 0.017 | 0.686 |
|  | Sen. | 0.500 | Sen. | 0.321 | Sen. | 0.179 |  |
| RF | PPV | **0.022** | PPV | **0.033** | PPV | **0.043** | **0.800** |
|  | Sen. | **0.643** | Sen. | **0.518** | Sen. | **0.321** |  |
| LGBM | PPV | 0.018 | PPV | 0.028 | PPV | 0.037 | 0.741 |
|  | Sen. | 0.518 | Sen. | 0.429 | Sen. | 0.286 |  |
| MLP | PPV | 0.013 | PPV | 0.019 | PPV | 0.020 | 0.638 |
|  | Sen. | 0.411 | Sen. | 0.286 | Sen. | 0.161 |  |
| W&D | PPV | 0.014 | PPV | 0.021 | PPV | 0.021 | 0.667 |
|  | Sen. | 0.464 | Sen. | 0.357 | Sen. | 0.214 |  |
| TabNet | PPV | 0.017 | PPV | 0.025 | PPV | 0.025 | 0.691 |
|  | Sen. | 0.500 | Sen. | 0.357 | Sen. | 0.179 |  |
| 3 years prediction window (prevalence = 0.89%) | | | | | | | |
| Model | 80% spec. | | 90% spec. | | 95% spec. | | AUROC |
| LogReg | PPV | 0.020 | PPV | 0.020 | PPV | 0.027 | 0.693 |
|  | Sen. | 0.446 | Sen. | 0.246 | Sen. | 0.154 |  |
| NBC | PPV | 0.020 | PPV | 0.021 | PPV | 0.020 | 0.673 |
|  | Sen. | 0.477 | Sen. | 0.246 | Sen. | 0.108 |  |
| RF | PPV | **0.024** | PPV | **0.028** | PPV | **0.036** | **0.757** |
|  | Sen. | **0.538** | Sen. | **0.323** | Sen. | **0.215** |  |
| LGBM | PPV | 0.020 | PPV | 0.025 | PPV | 0.026 | 0.747 |
|  | Sen. | 0.462 | Sen. | 0.292 | Sen. | 0.154 |  |
| MLP | PPV | 0.014 | PPV | 0.019 | PPV | 0.020 | 0.644 |
|  | Sen. | 0.323 | Sen. | 0.215 | Sen. | 0.123 |  |
| W&D | PPV | 0.018 | PPV | 0.020 | PPV | 0.022 | 0.679 |
|  | Sen. | 0.415 | Sen. | 0.277 | Sen. | 0.169 |  |
| TabNet | PPV | 0.022 | PPV | 0.024 | PPV | 0.028 | 0.704 |
|  | Sen. | 0.492 | Sen. | 0.292 | Sen. | 0.169 |  |

##### **Supplementary Figure 1.** Full data processing and cohort building chart flow, showing the steps were taken to go from the original RPDR database to the landmark-sampled data sets. NP: number of patients. NTV: number of total visits. NSV: number of sampled visits. NBD: number of patients who later developed early-onset BD. Note that since we defined the onset of BD between age 10 (inclusive) and 25 (non-inclusive) as the early onset, we required each patient to have at least 1 visit with the addition of the prediction window does not reach the 25^th^ birthday mark.

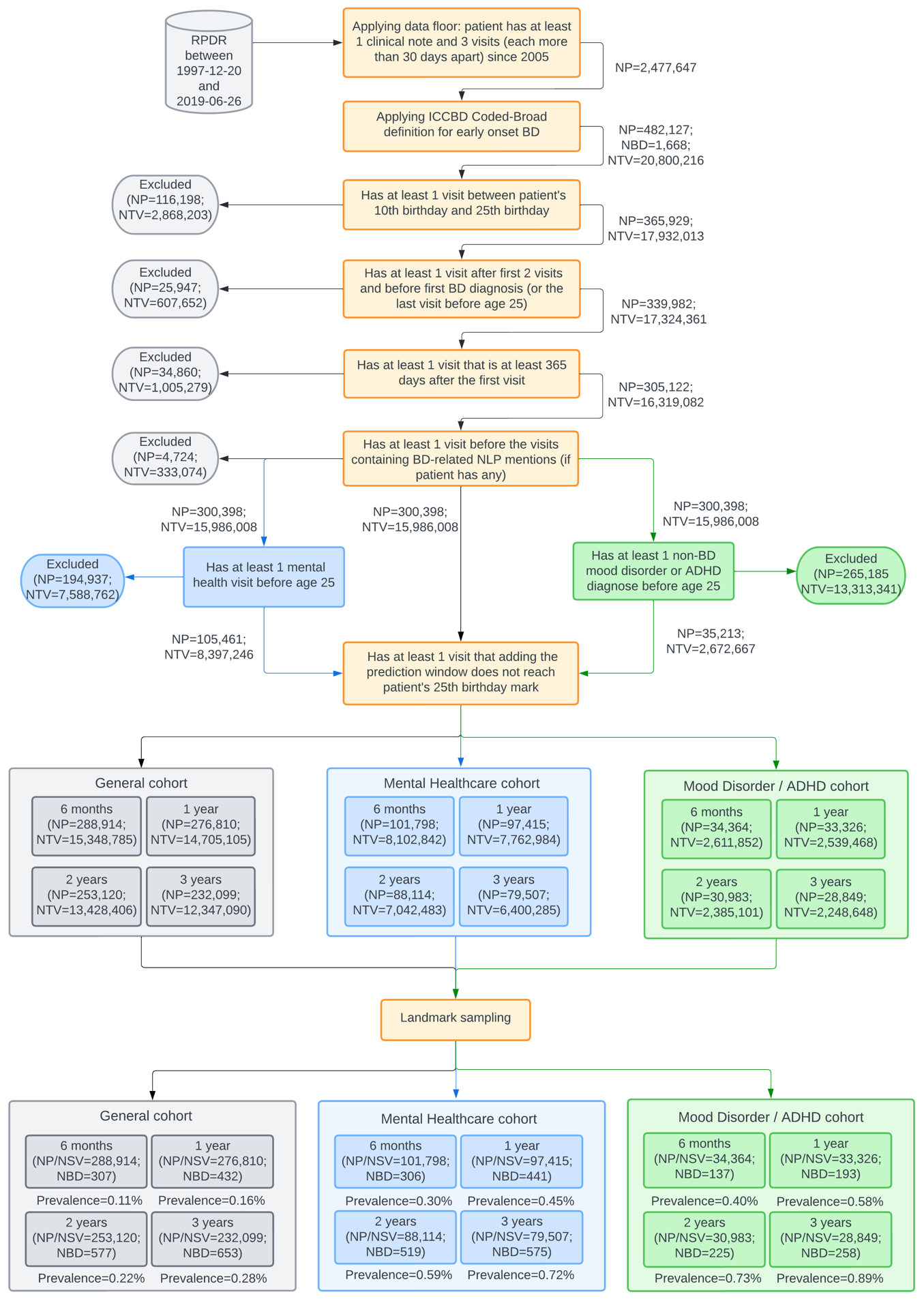

##### **Supplementary Figure 2.** Cumulative gain charts for all three cohort groups and four prediction windows. The results are from our Random Forest models. For each sub-plot, the percentage of patients in the test set (x-axis) ranked by the prediction probabilities (from left to right, i.e., from highest-risk quintile to lowest-risk quintile) is plotted against the percentage of early onset of bipolar disorder cases identified in the test set (y-axis). The color of each curve denotes the cohort group (blue: general cohort; green: mental healthcare cohort; orange: mood disorder/ADHD cohort).

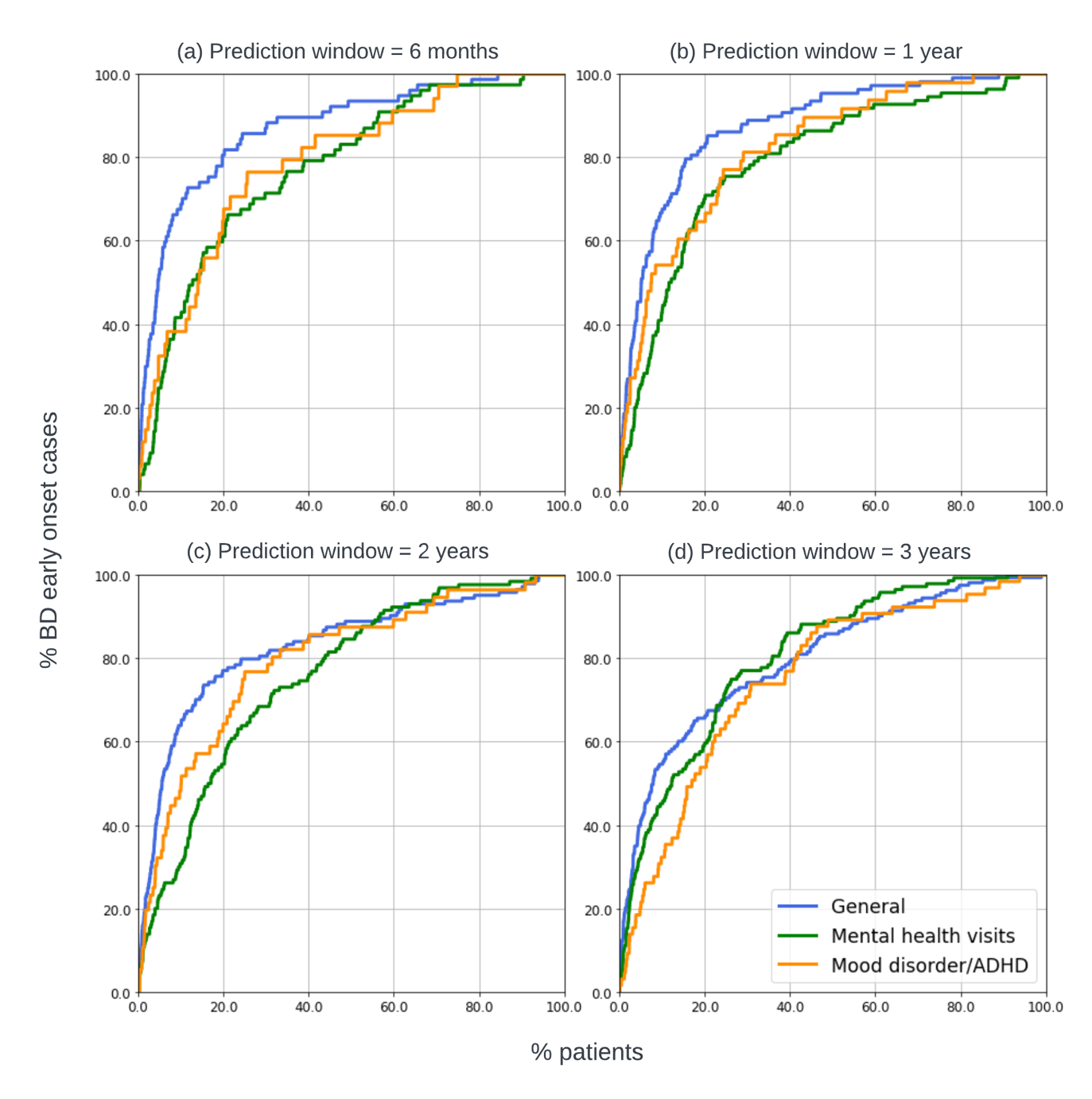

##### **Supplementary Figure 3.** Model metrics of RF-full using 2-year prediction window, stratified by patients’ age at the landmark visit.

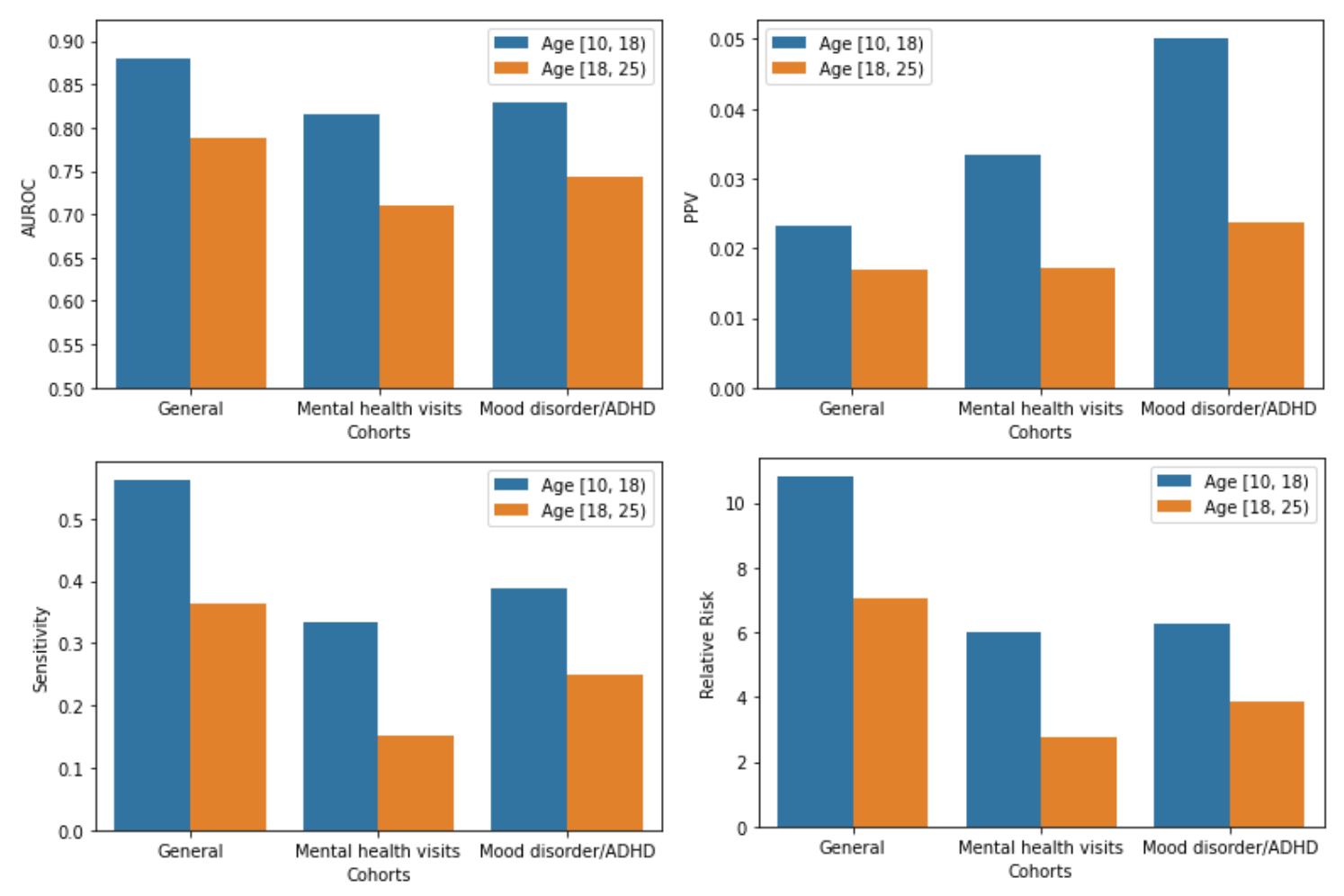

##### **Supplementary Figure 4.** Results using 2-year prediction window, stratified by number of visits each patient had before their “landmark visits”.

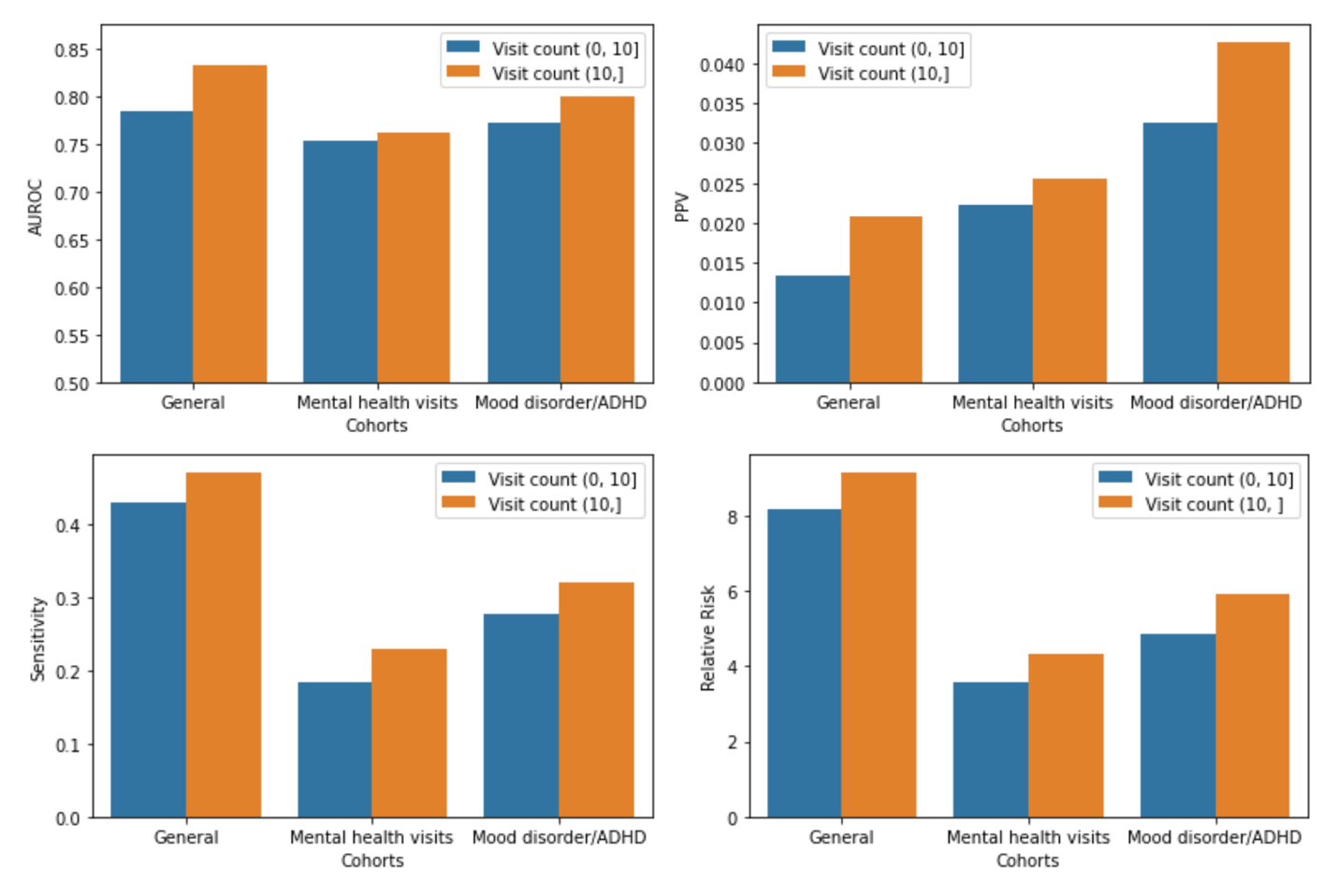

##### **Supplementary Figure 5.** Results using 2-year prediction window, stratified by gender (i.e., male vs. female).

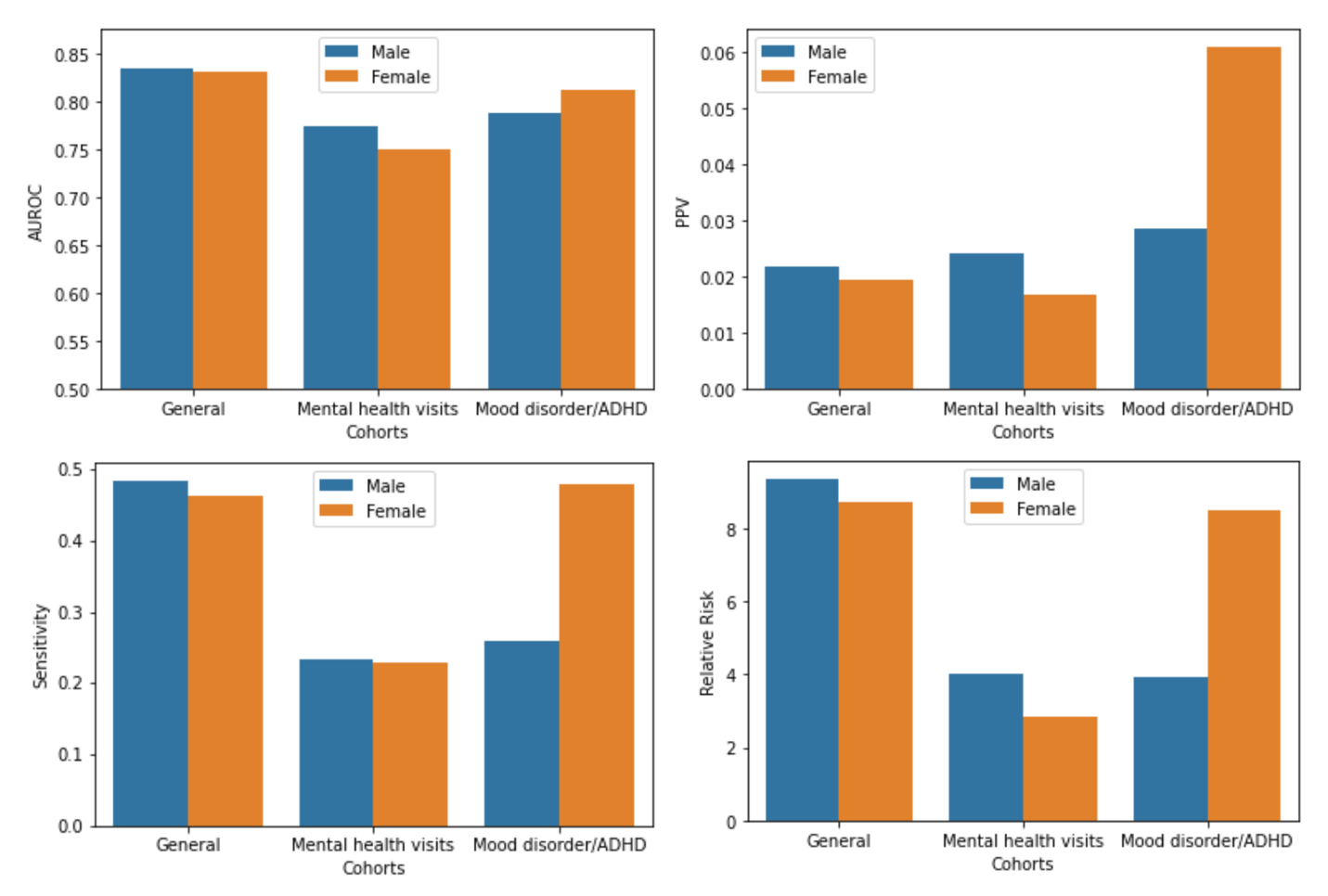

##### **Supplementary Figure 6.** Results using 2-year prediction window, stratified by race (i.e., white vs. non-white).

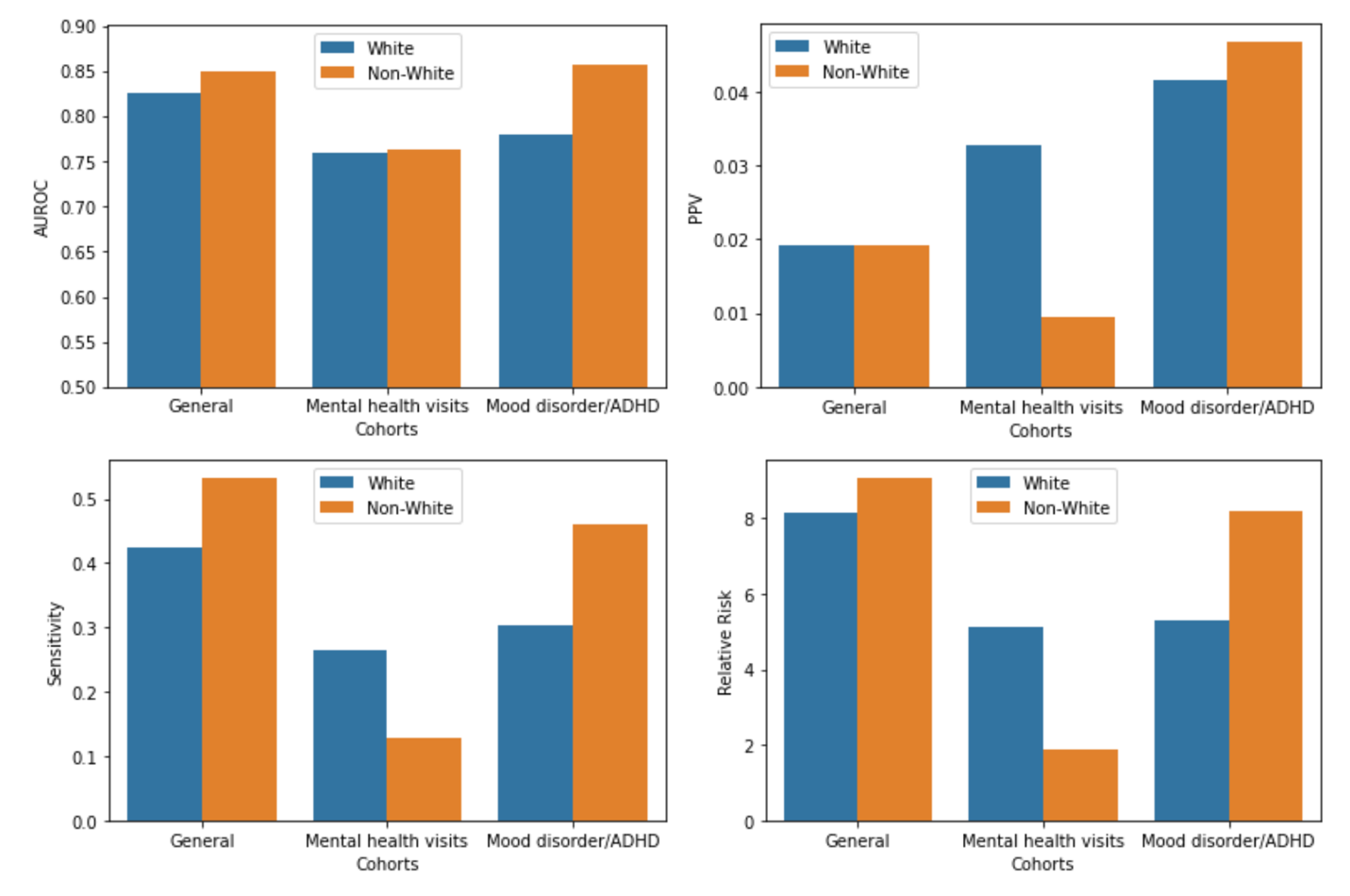

##### **Supplementary Figure 7 (a).** SHAP summary plots of the top-10 features for the General patients cohort with 6 months prediction window.

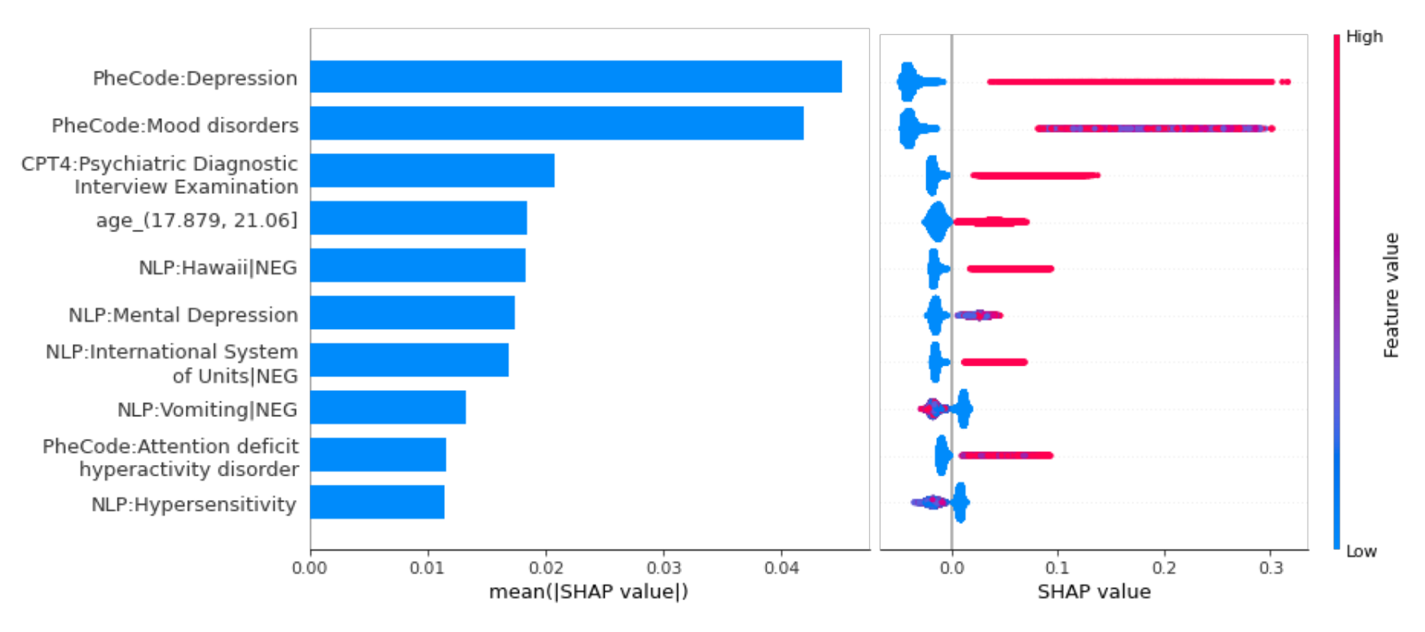

##### **Supplementary Figure 7 (b).** SHAP summary plots of the top-10 features for the Mental healthcare cohort with 6 months prediction window.

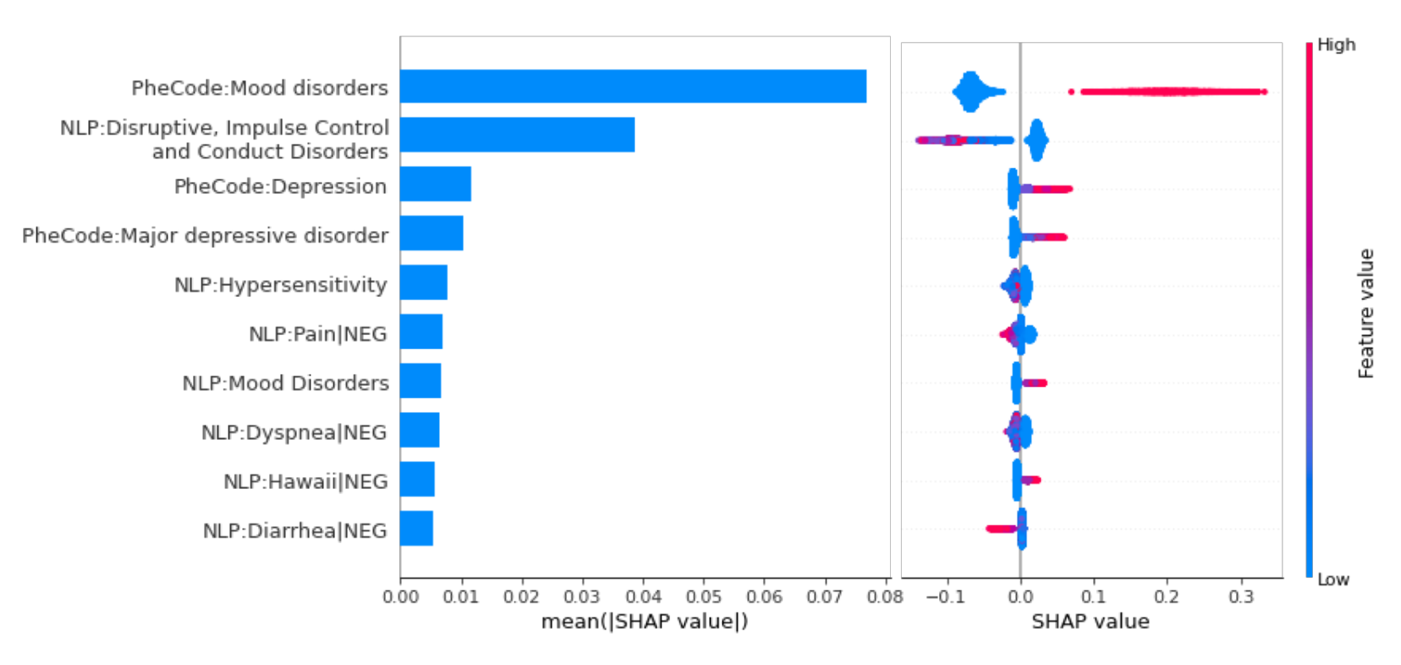

##### **Supplementary Figure 7 (c).** SHAP summary plots of the top-10 features for the Mood disorder/ADHD cohort with 6 months prediction window.

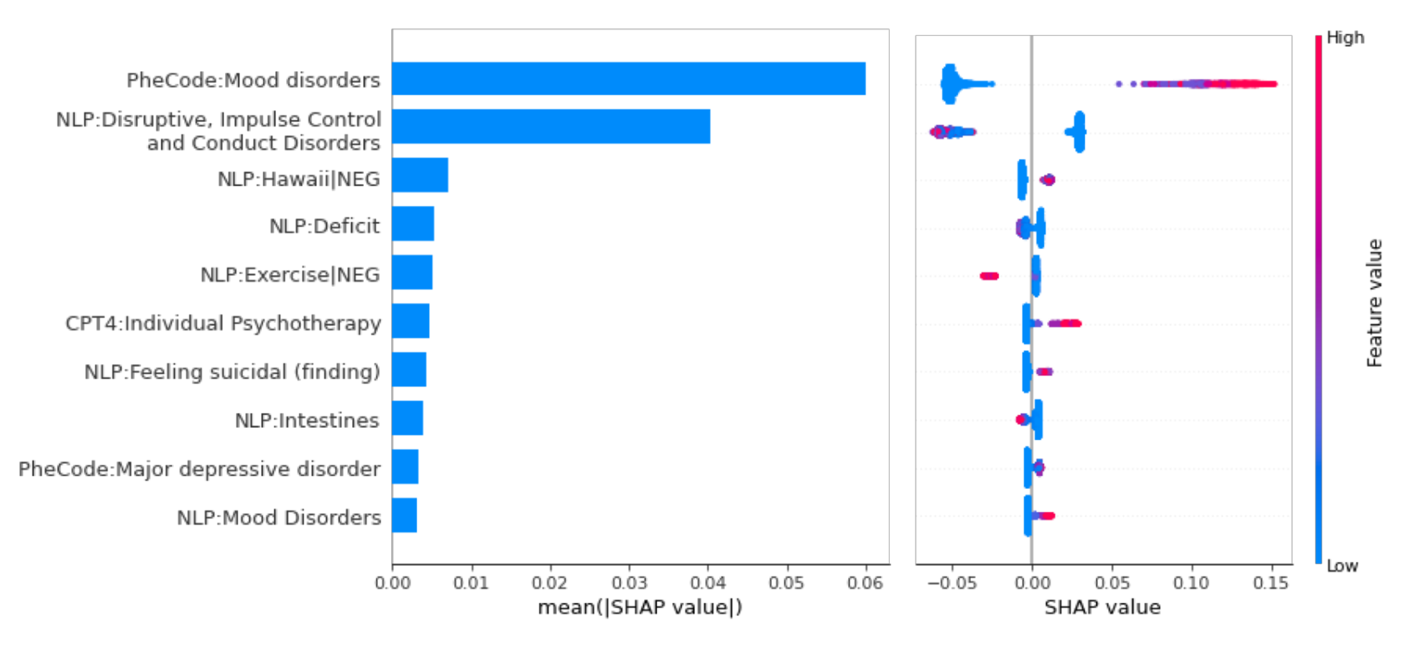

##### **Supplementary Figure 8 (a).** SHAP summary plots of the top-10 features for the General patients cohort with 1-year prediction window.

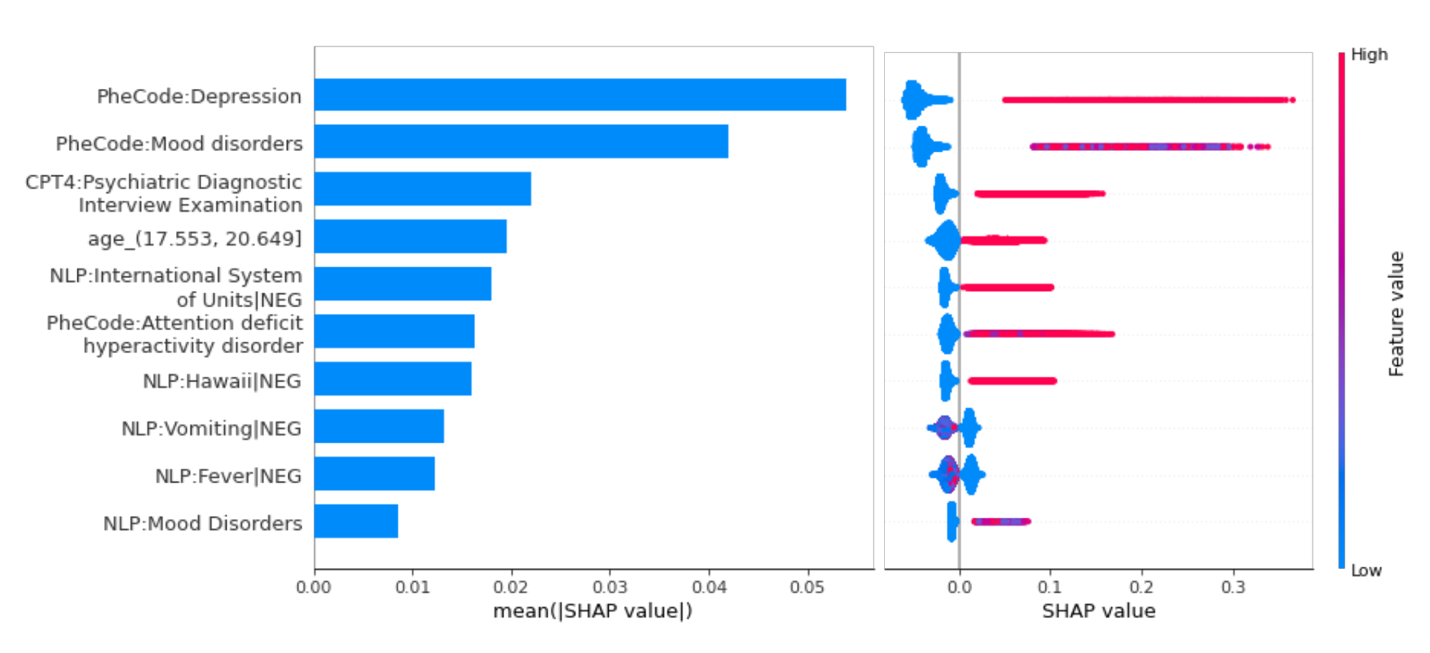

##### **Supplementary Figure 8 (b).** SHAP summary plots of the top-10 features for the Mental healthcare cohort with 1-year prediction window.

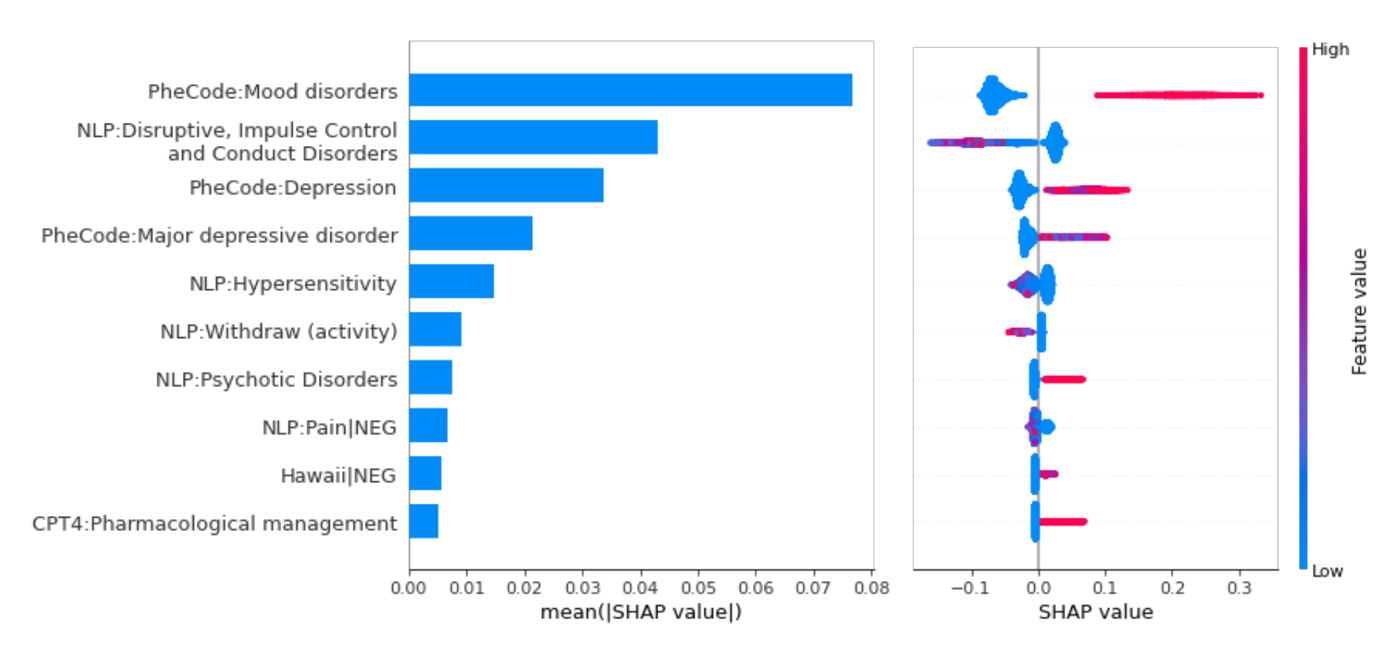

##### **Supplementary Figure 8 (c).** SHAP summary plots of the top-10 features for the Mood disorder/ADHD cohort with 1-year prediction window.

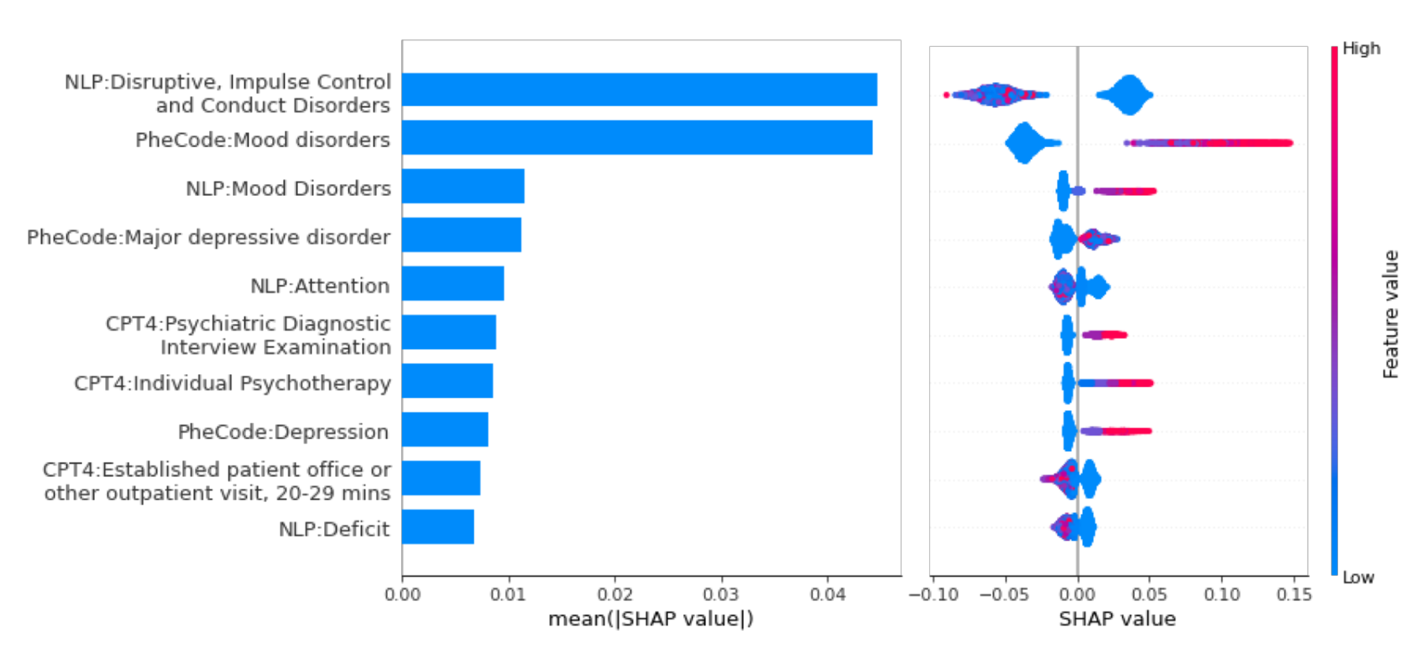

##### **Supplementary Figure 9 (a).** SHAP summary plots of the top-10 features for the General patients cohort with 3-year prediction window.

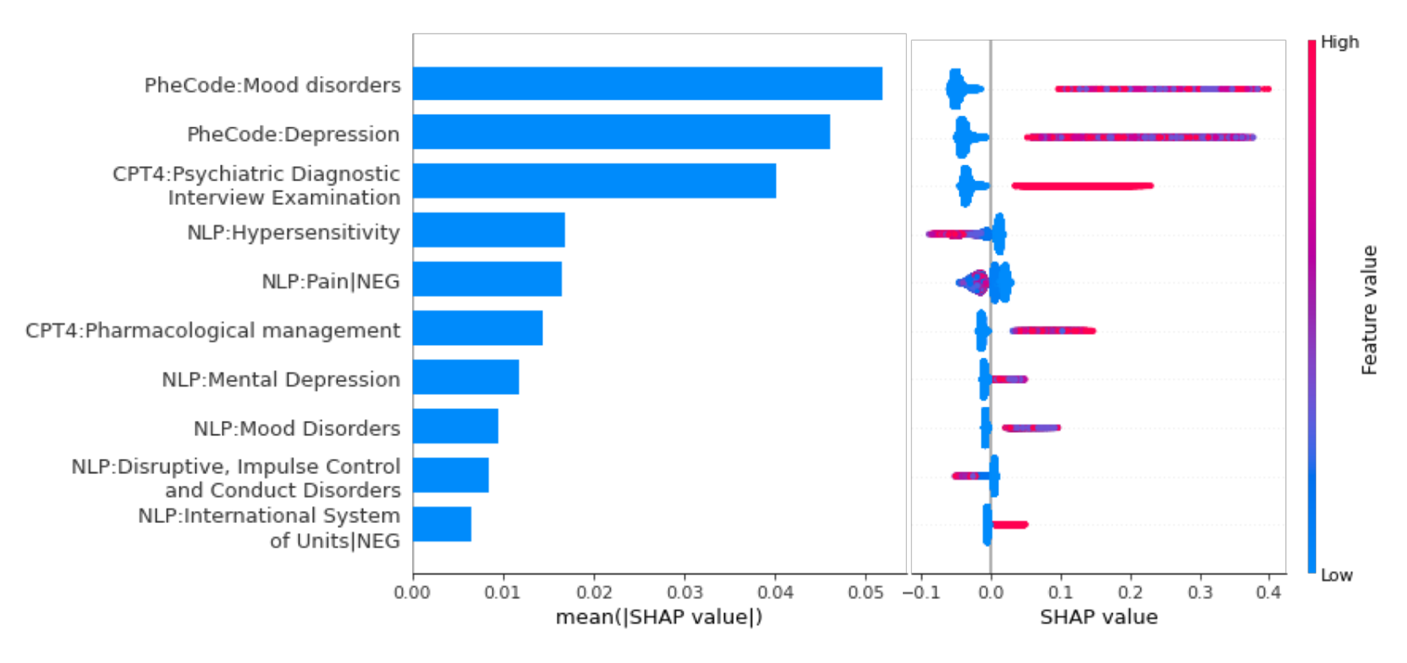

##### **Supplementary Figure 9 (b).** SHAP summary plots of the top-10 features for the Mental healthcare cohort with 3-year prediction window.

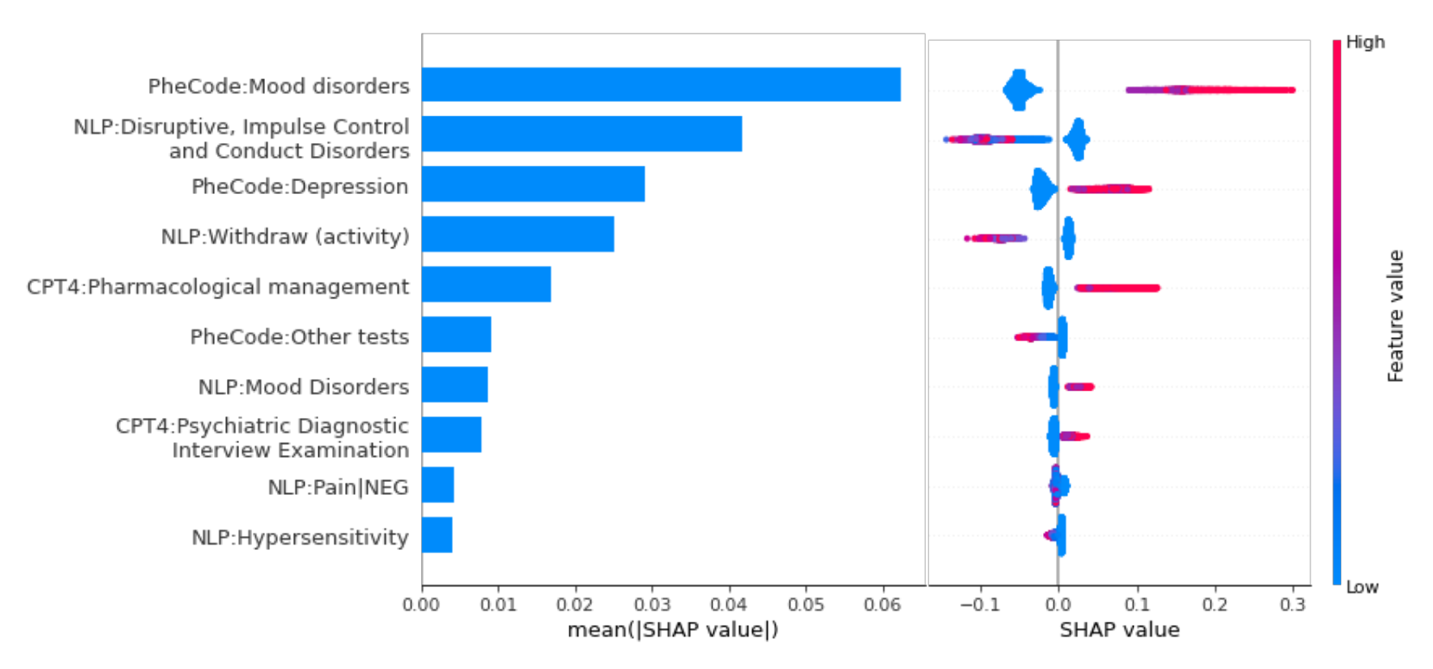

##### **Supplementary Figure 9 (c).** SHAP summary plots of the top-10 features for the Mood disorder/ADHD cohort with 3-year prediction window.

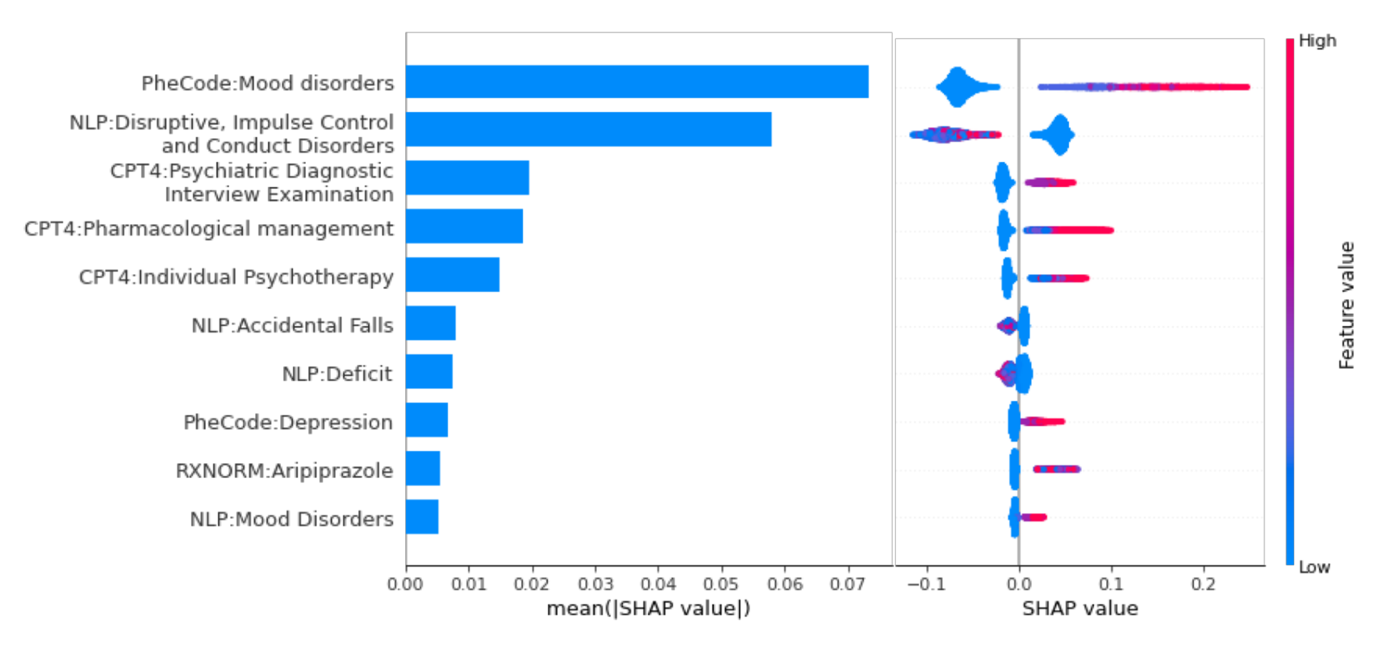

##### **Supplementary Figure 10.** Model performance comparison between the original RF model and the light-weight models, with 6 months prediction window.

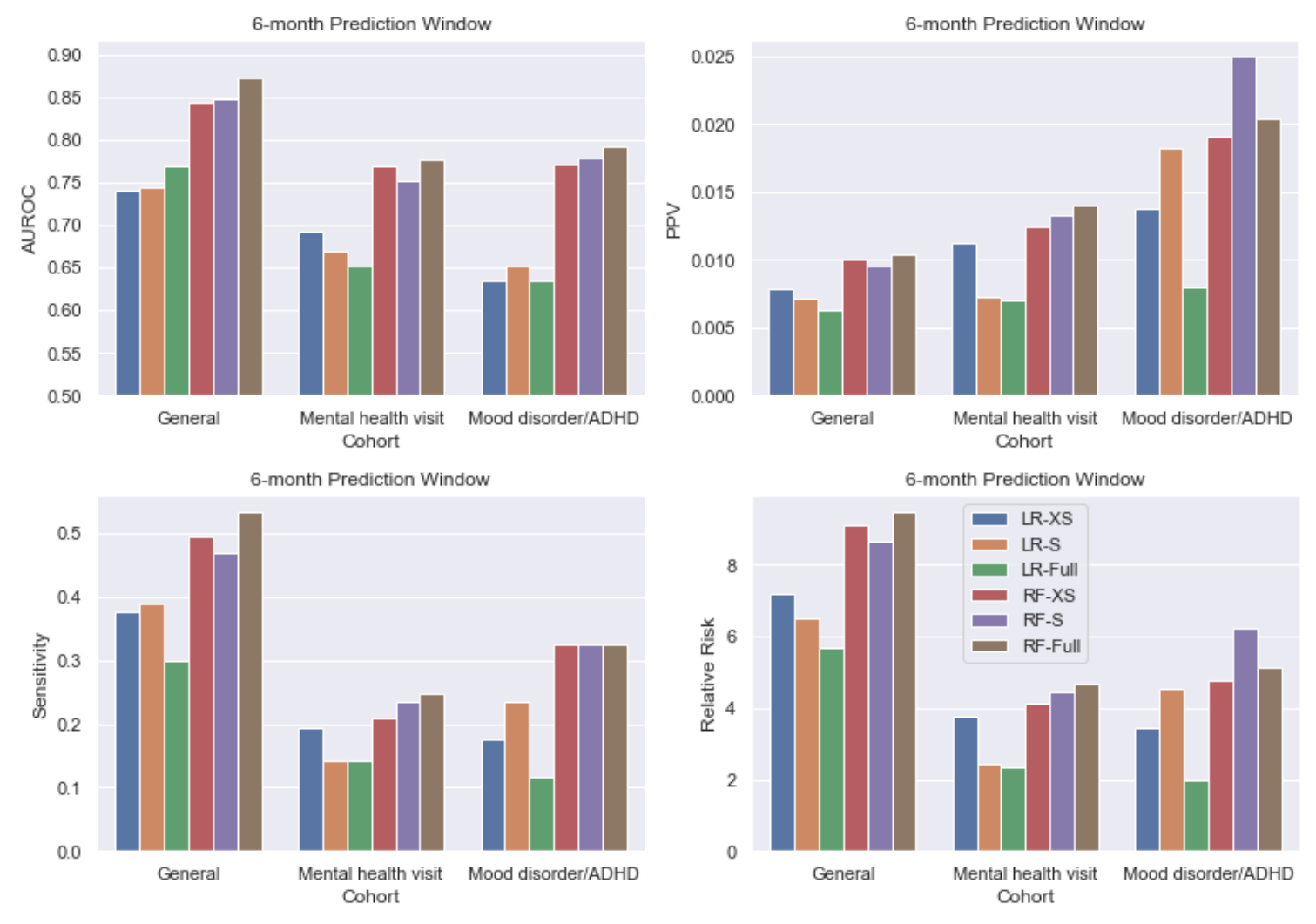

##### **Supplementary Figure 11.** Model performance comparison between the original RF model and the light-weight models, with 1-year prediction window.

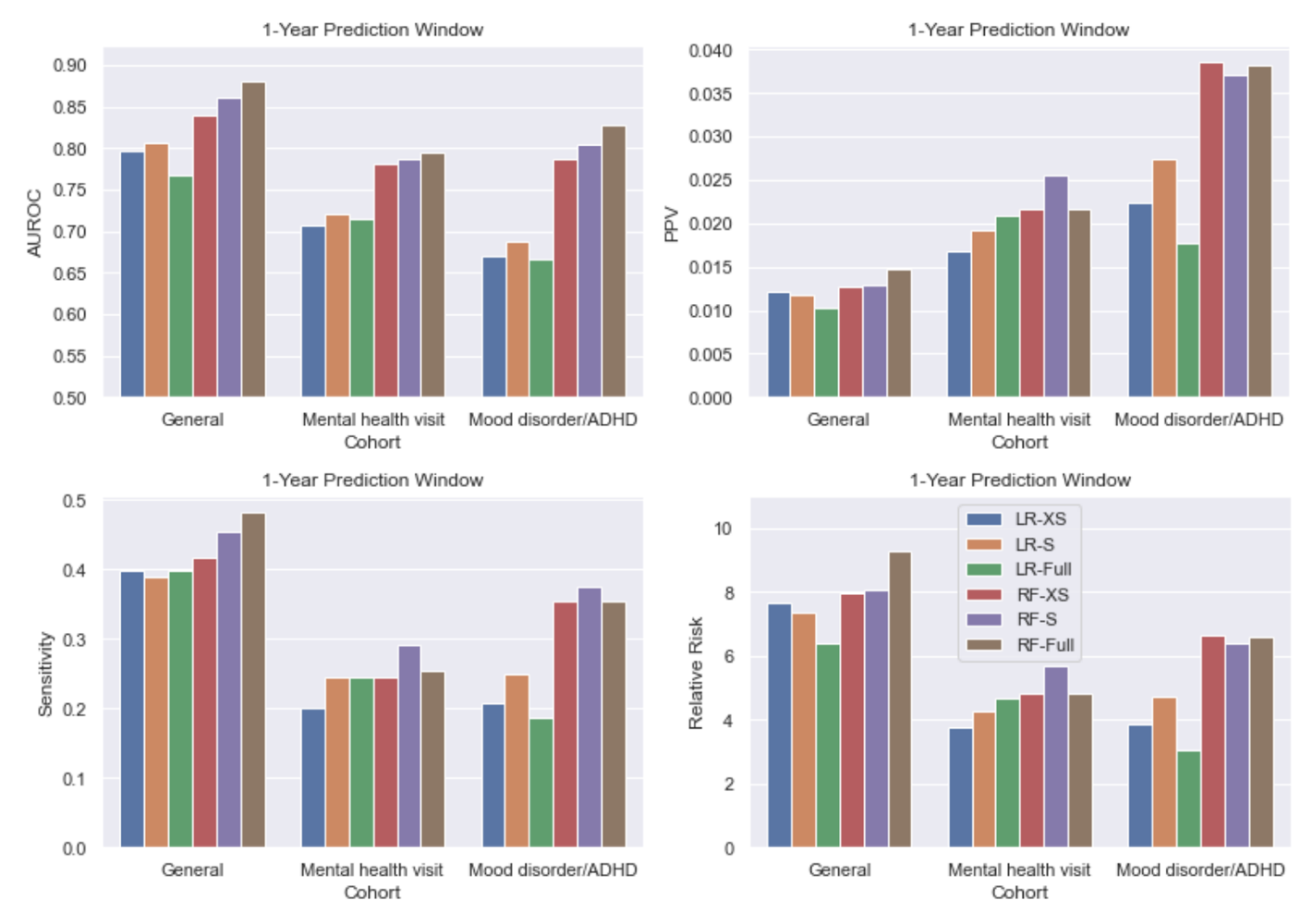

##### **Supplementary Figure 12.** Model performance comparison between the original RF model and the light-weight models, with 3-year prediction window.

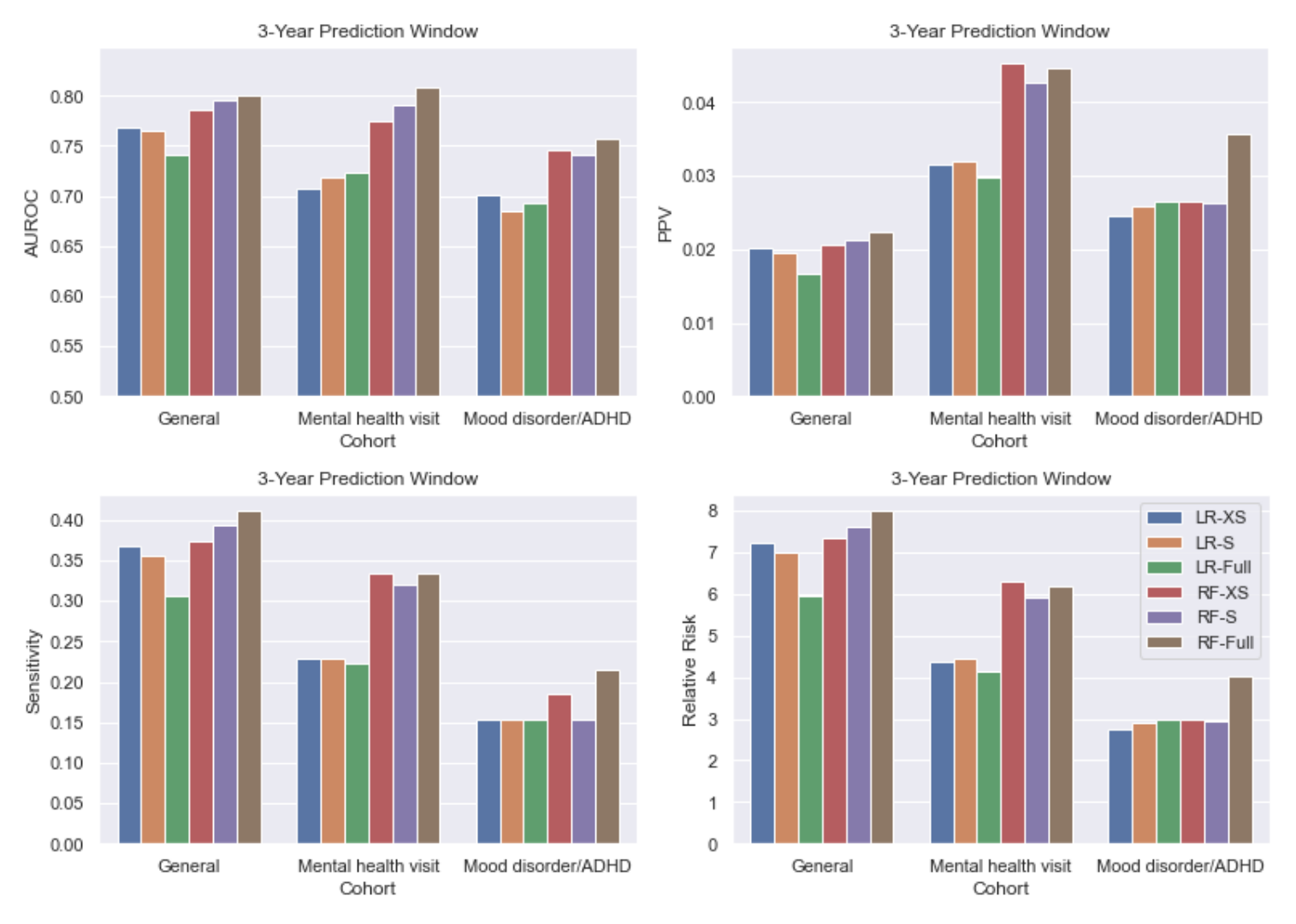
